# A single blinded, phase IV, adaptive randomised control trial to evaluate the safety of coadministration of seasonal influenza and COVID-19 vaccines (The FluVID study)

**DOI:** 10.1101/2023.06.14.23291380

**Authors:** JA Ramsay, M Jones, AM Vande More, SL Hunt, PCM Williams, M Messer, N Wood, K Macartney, FJ Lee, WJ Britton, TL Snelling, ID Caterson

**Affiliations:** Telethon Kids Institute, PO Box 855, West Perth, Western Australia, 6872. Australia; School of Public Health, University of Sydney. NSW 2006 Australia; Royal Prince Alfred Hospital, Camperdown. 2050 NSW Australia; National Centre for Immunisation Research and Surveillance, Westmead, New South Wales, Australia; The Children’s Hospital at Westmead Clinical School, University of Sydney, NSW, 2006, Australia; Department of Clinical Immunology & Allergy, Royal Prince Alfred Hospital, Camperdown, NSW 2050, and Sydney Medical School, University of Sydney, Sydney, Australia; Centenary Institute, University of Sydney. NSW 2006 Australia; Department of Endocrinology, Royal Prince Alfred Hospital, Camperdown, NSW 2050, and Boden Initiative, Charles Perkins Centre, University of Sydney. NSW 2006 Australia

**Keywords:** Coadministration, COVID-19 vaccination, Influenza vaccination, Adverse events, Safety

## Abstract

**Objectives:** We evaluated the frequency of moderate and severe adverse events following coadministration of seasonal influenza vaccine (SIV) versus placebo with COVID-19 vaccines among adults to support practice guidelines.

**Methods:** FluVID is a participant-blinded, phase IV, randomised control trial. On the same day as the participant’s scheduled COVID-19 vaccine, participants were randomised to receive SIV or saline placebo; those assigned placebo at visit one then received SIV a week later, and vice versa. Self-reported adverse events were collected for daily seven days following each visit.

The primary endpoint was any solicited adverse event of at least moderate severity occurring up to seven days following receipt of SIV or placebo. This was modelled using a Bayesian logistic regression model. Analyses were performed by COVID-19 vaccine type and dose number.

**Results:** Overall, 248 participants were enrolled; of these, 195 had received BNT162b2 and 53 had received mRNA1273 COVID-19 vaccines according to national guidelines. After randomisation, 119 were assigned to receive SIV and 129 were assigned to receive placebo at visit one.

Adverse events were most frequently reported as mild (grade 1) in nature. Among 142 BNT162b2 booster dose one and 43 BNT162b2 booster dose two recipients, the posterior median risk difference for moderate/severe adverse events following SIV versus placebo was 13% (95% credible interval [CrI] -0.03 to 0.27) and 13% (95%CrI -0.37 to 0.12), respectively. Among 18 mRNA1273 booster dose one and 35 mRNA1273 booster dose two recipients, the posterior median risk difference of moderate/severe adverse events following influenza vaccine versus placebo was 6% (95%CrI -0.29 to 0.41) and -4% (95%CrI -0.30 to 0.23), respectively.

**Conclusion:** Adverse events following SIV and COVID-19 co-administration were generally mild and occurred with similar frequency to events following COVID-19 vaccine alone. We found no evidence to justify routine separation of SIV and COVID-19 vaccine doses.

**Clinical trial registration:** ACTRN12621001063808

**Highlights:**

- The coadministration of mRNA COVID-19 and influenza vaccines typically resulted in mild events that were limited to 4 days.
- Frequency and nature of adverse events were similar to those in other randomised trials.
- This trial demonstrates a suitable design for evaluating vaccine schedules and coadministration.

## Introduction

In October 2021, the World Health Organization developed guidance for the coadministration of seasonal influenza vaccines (SIV) and vaccines to prevent against coronavirus diseases 2019 (COVID-19) noting that coadministration of these vaccines was already occurring in many countries despite a very limited evidence base pertaining to this practice (1). Despite this guidance, concerns remained around the potential for increased reactogenicity of these vaccines if administered at a single visit.

If safe to do so, co-administering scheduled vaccines at a single visit is usually convenient and may reduce the risk of delayed or missed doses. Many vaccine providers and members of the public were initially hesitant toward coadministration of COVID-19 and SIV (2,3) given the paucity of the evidence of the safety of coadministration. Subsequently, evidence of acceptable safety and immunogenicity gradually emerged (4,5), and many authorities now permit or recommend coadministration of COVID-19 and influenza vaccines (6,7). However, despite the continual emergence of new evidence to support co-administration, practice remains limited.

Several studies have shown varying immune responses when assessing the coadministration of COVID-19 and influenza vaccines, considering differences study designs, populations and COVID-19 vaccine types and doses. Yet, safety outcomes have generally been comparable across these studies (4,5,8,9). Trials investigating simultaneous administration of ChAdOx1 or BNT162b2 vaccine, NVX-CoV2373, and mRNA-1273 alongside an age-appropriate influenza vaccine demonstrated similar safety, reactogenicity and antibody responses. However, both the NVX-CoV2373 and mRNA-1273 studies were open label and not placebo controlled, limiting differentiation of the true effect. A randomised and controlled study of adults ≥ 60 years in the Netherlands, who received BNT162b2 and a non-adjuvanted influenza vaccine found coadministration to be safe, although the functionality and magnitude of antibody response to SARS-CoV-2 was reduced (9). Further evidence is required to fully understand the potential interactions between these vaccines.

Here we present the results of a randomised controlled trial to evaluate the reactogenicity caused by coadministration of SIV and COVID-19 vaccines.

## Methods

### Study design and participants

FluVID was a participant-blinded, phase IV, Bayesian sequential multi-arm placebo-controlled trial with an adaptive sample size; it incorporated pre-specified stopping rules for both inferiority and non-inferiority. The trial was pragmatic and conducted across two community COVID-19 vaccination centres in Sydney, Australia. The maximum sample size was set to 1,000 participants based on feasibility considerations including available funding and resources. Analyses were planned to commence after enrolment and 7-day follow-up of 200 participants who had received any one of the available COVID-19 vaccines, and then after every subsequent 100 enrolments. Data collection and endpoints were harmonised with the ComFluCov study in the United Kingdom (UK) (4).

Participants were adults (≥ 18 years old) who had received an approved COVID-19 vaccine through the national COVID-19 vaccination program on the day of enrolment, and who were also eligible for SIV per Australian guidelines. People who were severely immunocompromised or who had received SIV within the previous six months were excluded.

Potential participants were approached while awaiting their COVID-19 vaccine dose at the vaccination centre; they were able to pre-register their interest by responding to text messages sent prior to their vaccination appointment or to electronic advertisements distributed to local health staff. All participants provided written informed consent.

### Vaccines

The SIVs used were based on national age-based recommendations and determined by their availability at the sites. Adults <65 years old received either Fluarix Tetra (GlaxoSmithKline) or FluQuadri (Sanofi) which are both non-adjuvanted, inactivated quadrivalent SIVs containing 60 μg of haemagglutinin (HA). Adults ≥ 65 years old received Fluad Quad (Seqirus), an adjuvanted quadrivalent SIV containing 60 μg of HA. The placebo used 0.5 ml of intramuscular 0.9% sodium chloride.

Over the study period, the COVID-19 vaccines delivered to participants through the national COVID-19 vaccination program were BNT162b2 (Pfizer-BioNTech) as a primary series or third (first booster) or fourth (second booster) COVID-19 vaccine dose; and mRNA1273 (Moderna) as a third (first booster) or fourth (second booster) COVID-19 vaccine dose.

### Procedures

At visit one, participants were randomly assigned to receive either a SIV or a saline placebo within six hours of receiving their COVID-19 vaccine dose. An unblinded study nurse administered the applicable vaccine based on a computer-generated randomisation stored and assigned via the electronic data-capture system. The trial statistician provided the randomisation list for the two treatment groups stratified by site, COVID-19 vaccine type and dose number received, and SIV type. The list was computer-generated using random permuted blocks of sizes 2 to 10. The allocation ratio within each stratum was 1:1.

The unblinded study nurse administered the influenza vaccine or saline placebo in the opposite arm to that of the participant’s COVID-19 vaccine dose. SIV and placebo were prepared in non-identical, pre-filled syringes which were covered with a label and concealed within an opaque box until ready for administration. SIV and placebo were delivered intramuscularly by authorised nurse immunisers in accordance with the Australian guidelines. Participants were observed for at least 15 minutes afterwards. Participant blinding was maintained by asking the vaccination recipient to look away while the SIV or placebo was delivered intramuscularly into the upper arm. Additionally, updating the participant electronic immunisation record was delayed until completion of the safety follow-up.

Seven to fourteen days after visit one, participants received either a saline placebo or SIV if they had been assigned to receive SIV or placebo at visit one, respectively.

### Assessment of Outcomes

Research staff involved in the safety and reactogenicity follow-up were blinded to the group assignment of participants. All study data were collected and managed using REDCap electronic data capture tools hosted at Sydney Local Health District (10,11). Participants were provided with a thermometer and tape measure and were instructed to record their temperature, as well as measure any swelling or erythema at the injection site at the same time each day. Local adverse events (injection site pain, swelling, and erythema) and systemic adverse events (temperature, headache, fatigue, chills, myalgia, joint pain, nausea and vomiting, and diarrhoea) were solicited via a daily electronic diary card sent via SMS or email for 7 days after each study visit. Outcomes were self-graded as absent (0), mild (1), moderate (2), severe (3), or potentially life threatening (4). Grading aligned with the FDA’s Toxicity Grading Scale (12). Samples of the surveys provided can be found in appendix B. Any serious adverse events (SAEs), pre-specified adverse events of special interest (AESIs), medical attendances and days absent from work up to 21 days after enrolment were collected (described in further detail in appendix A and C).

### Statistical analysis

FluVID was designed assuming a maximum sample size of 1,000 participants due to feasibility and resource constraints. Subsequent interim analyses were planned for every additional 100 participants. With inclusion of two COVID-19 vaccine types (brands) and based on a range of possible effect sizes, trial simulations depicted a median sample size of 700. Further details are provided in the statistical analysis plan (appendix D).

The primary endpoint was any recorded adverse event of at least moderate severity (grade ≥ 2) occurring up to 7 days following receipt of SIV or placebo. This was modelled using a Bayesian logistic regression model. Covariates included age (decadal), sex, and relevant comorbidities (type 2 diabetes, hypertension, and heart disease) as well as COVID-19 vaccine type and dose number received, and SIV type. Coadministration of a SIV with a COVID-19 vaccine was pre-defined as having non-inferior reactogenicity to coadministration of saline placebo with a COVID-19 vaccine if the probability of a <15% increased risk of grade ≥ 2 adverse events was greater than 0.985.

Analyses were performed using both the intention-to-treat (ITT) and per-protocol (PP) populations. The ITT population included all those randomised in accordance with their assigned group, irrespective of any protocol deviations. The PP population included all those randomised who: were deemed eligible to participate, received the correct age-appropriate SIV or placebo within 6 hours of their COVID-19 vaccine dose, provided responses to ≥ 90% of the survey questions on all 7 daily diary cards, and who did not withdraw prior to ascertainment of the primary endpoint.

### Trial ethics and role of the funding source

FluVID was supported by funding from Sydney Local Health District, Snow Medical Foundation, and the New South Wales Ministry of Health; none had any commercial interest in the study outcome none had any role in study design, data collection, data analysis, data interpretation, or writing of the report.

The trial was approved by the Sydney Local Health District (Royal Prince Alfred Hospital) Ethics Committee (2021/ETH01232) and was sponsored by Sydney Local Health District. The approved study protocol is included in appendix A.

## Results

The study stopped early due to challenging recruitment and slowed uptake of COVID-19 vaccine in the study population. Enrolment was suspended on 10th of October 2022, and a final analysis was conducted before reaching the first planned analysis trigger. Between 2nd November 2021 and 10th October 2022, 252 participants were enrolled, of these, 195 had received BNT162b2 (1 first dose, 3 second dose, 147 first booster, 44 second booster) and 53 participants had received mRNA1273 (18 first booster, 35 second booster). Of the 248 who were then randomised, 119 were assigned to receive an SIV and 129 were assigned to receive placebo at study visit 1 (Figure 1); 219 and 29 participants received a non-adjuvanted and adjuvanted SIV, respectively

**Figure 1.**
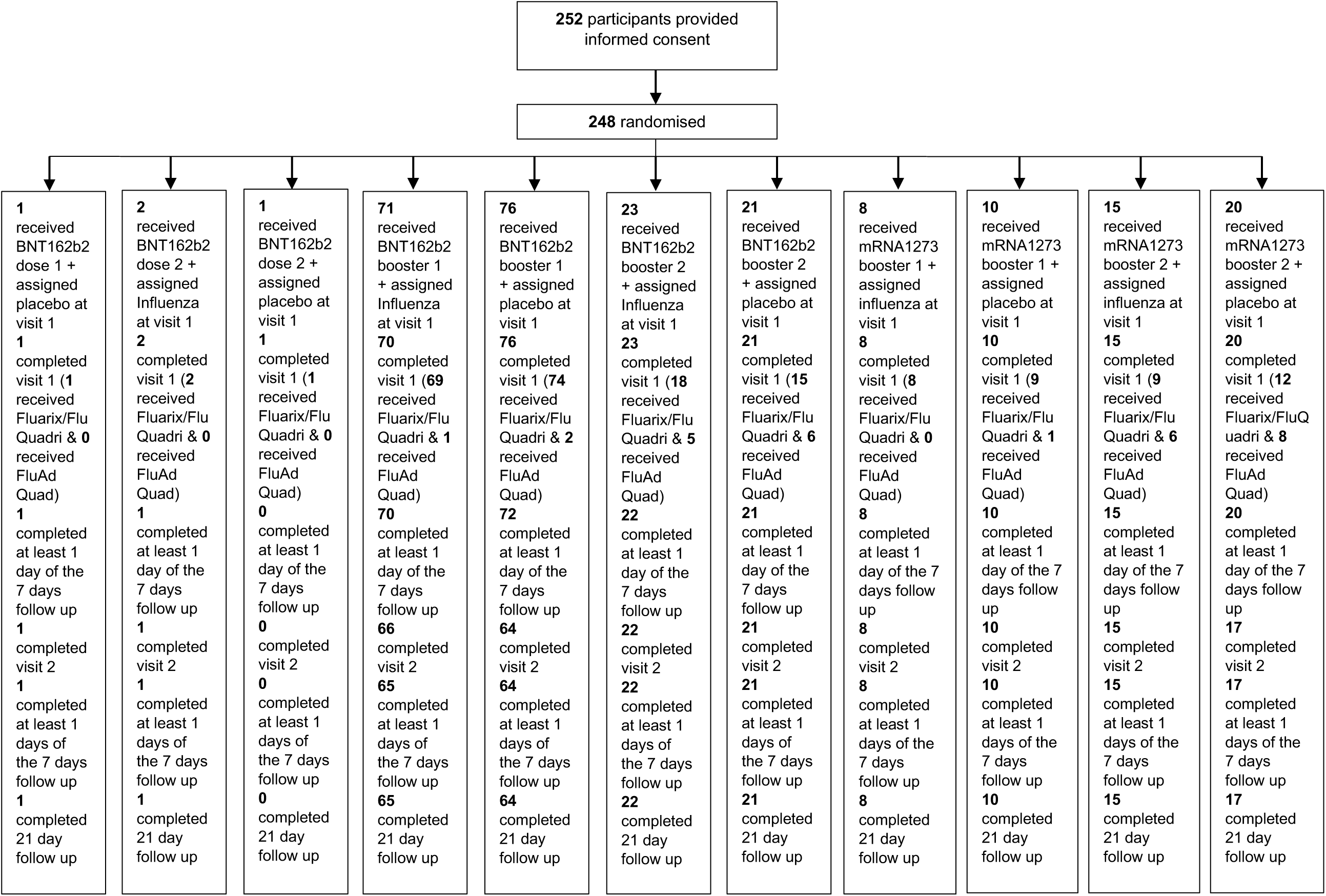
Participant disposition: 252 participants provided informed consent and 248 participants were randomised (2 participants were found to be ineligible after consent was provided). One participant in the who received BNT162b2 booster 1 was randomised but did not receive the placebo they were randomised to. 18 participants were lost to follow up and 4 participants withdrew.

Among the 248 randomised participants, 241 (97%) were included in the ITT analysis (117 in SIV and 124 assigned in placebo at visit one), and 230 (93%) were included in the PP analysis (112 who received SIV and 118 who received placebo at visit one) (Figure 1). Of those who completed visit one, 18 did not return for visit two (six assigned to receive SIV and 12 assigned to receive placebo at visit 1). The day 21 follow-up survey was completed by 225 (90%) randomised participants. Two participants were deemed ineligible after randomisation due to previous receipt of an influenza vaccine and were therefore excluded from the PP analysis.

The baseline characteristics between those assigned to SIV versus placebo at visit one were similar (table 1). The median age of participants was 42.4 years (IQR 31.4 – 55.6); 128 were male (52%) and 181 (73%) were of European/Caucasian ethnicity. The median BMI was 25.1 (IQR 22.8, 27.8) and 222 (90%) reported no comorbidities

**Table 1.** Participant demographics and baseline characteristics

|  |  | BNT162b2 booster 1 |  | BNT162b2 booster 2 |  | mRNA1273 booster 1 |  | mRNA1273 booster 2 |  |
| --- | --- | --- | --- | --- | --- | --- | --- | --- | --- |
|  |  | Intervention | Placebo | Intervention | Placebo | Intervention | Placebo | Intervention | Placebo |
|  |  | N = 71 | N = 76 | N = 23 | N = 21 | N = 8 | N = 10 | N = 15 | N = 20 |
| **Age** |  | 36.3<br>(28.0,48.5) | 36.2<br>(28.9,47.1) | 51.4<br>(44.2,59.2) | 50.6<br>(43.0,61.4) | 35.6<br>(33.6,50.6) | 37.5<br>(29.6,43.8) | 59.1<br>(56.3,67.7) | 59.8<br>(51.7,65.5) |
| **Sex** | Male | 31 (44) | 37 (49) | 16 (70) | 8 (38) | 5 (62) | 4 (40) | 10 (67) | 14 (70) |
|  | Missing | - | 1 (1) | - | - | - | - | - | - |
| **BMI** |  | 24.4<br>(22.8,27.0) | 24.9<br>(22.3,29.3) | 26.2<br>(24.1,27.9) | 26.0<br>(24.8,27.5) | 24.5<br>(20.6,26.6) | 25.7<br>(22.7,27.1) | 25.1<br>(23.9,27.7) | 25.5<br>(23.0,27.4) |
| **Smoking status** |  |  |  |  |  |  |  |  |  |
|  | Never | 52 (73) | 57 (75) | 18 (78) | 18 (86) | 8 (100) | 8 (80) | 11 (73) | 12 (60) |
|  | Former | 13 (18) | 10 (13) | 4 (17) | 2 (10) | - | 2 (20) | 4 (27) | 7 (35) |
|  | Current | 6 (8) | 8 (11) | 1 (4) | 1 (5) | - | - | - | 1 (5) |
|  | Missing | - | 1 (1) | - | - | - | - | - | - |
| **Ethnicity** | Asian | 7 (10) | 11 (14) | 2 (9) | 1 (5) | 2 (25) | 1 (10) | - | 2 (10) |
|  | Black African | - | - | - | - | - | - | - | - |
|  | European Caucasian | 56 (79) | 48 (63) | 17 (74) | 17 (81) | 4 (50) | 6 (60) | 14 (93) | 17 (85) |
|  | Indian subcontinent | 2 (3) | 6 (8) | 3 (13) | 1 (5) | 1 (12) | 2 (20) | - | 1 (5) |
|  | Indigenous Australia | - | - | - | - | - | - | - | - |
|  | Middle Eastern | 3 (4) | 3 (4) | - | - | - | 1 (10) | - | - |
|  | Pacific Islander | 1 (1) | 1 (1) | - | - | - | - | - | - |
|  | South American/<br>Spanish/local India descent | 1 (1) | 3 (4) | - | 2 (10) | 1 (12) | - | - | - |
|  | Missing | 1 (1) | 4 (5) | 1 (4) | - | - | - | 1 (7) | - |
| **Occupation** | Aged and disability care worker | - | 2 (3) | - | - | - | - | - | - |
|  | Border and quarantine staff | 1 (1) | 2 (3) | - | - | - | - | 1 (7) | - |
|  | Diagnostic laboratory staff | 1 (1) | - | - | - | - | - | - | - |
|  | Healthcare worker | 13 (18) | 16 (21) | 1 (4) | 2 (10) | - | 1 (10) | 2 (13) | 1 (5) |
|  | Missing | - | 1 (1) | - | - | - | - | - | - |
|  | Other | 56 (79) | 55 (72) | 22 (96) | 19 (90) | 8 (100) | 9 (90) | 12 (80) | 19 (95) |
| **Pregnancy status** | Yes | - | - | - | - | - | - | - | - |
|  | Missing | - | - | - | - | - | - | 1 (9) | - |
| **Vaccine received in the preceding 30 days** | Yes | 1 (1) | - | - | - | - | - | - | - |
|  | Missing | - | 1 (1) | - | - | - | - | - | - |
| **Comorbidities recorded** | Any comorbidity | 6 (17) | 2 (6) | 4 (29) | 4 (22) | 2 (40) | - | 3 (38) | 4 (25) |
|  | Allergies | 14 (40) | 15 (48) | 4 (29) | 4 (22) | 1 (20) | 4 (100) | 1 (13) | 3 (19) |
|  | Diabetes | 2 (6) | 1 (3) | 1 (7) | 1 (6) | 1 (20) | - | 1 (13) | 2 (13) |
|  | Cardiovascular disorder | 5 (14) | 2 (6) | 3 (21) | 3 (17) | 1 (20) | - | 2 (25) | 4 (25) |
|  | Respiratory disease | 4 (11) | 5 (16) | 1 (7) | 3 (17) | - | - | 1 (13) | 2 (13) |
|  | Immuno-compromising disorder | 2 (6) | 2 (6) | - | 1 (6) | - | - | - | - |
|  | Liver disease | 1 (3) | - | - | - | - | - | - | - |
|  | Renal disease | - | - | 1 (7) | - | - | - | - | 1 (6) |
|  | Cancer (current solid organ or haematological malignancy) | 1 (3) | 4 (13) | - | 2 (11) | - | - | - | - |
|  | Blood products in last 6 months | - | - | - | - | - | - | - | - |
| **On medication at enrolment** |  | 25 (35) | 27 (36) | 8 (35) | 5 (24) | 3 (38) | 1 (10) | 6 (40) | 7 (35) |

Electronic diary cards from day 1–7 after visit one were fully or partially completed by 241 (97%) participants allowing ascertainment of the primary endpoint. One or more grade ≥ 2 adverse events were self-reported by 67 (56%) participants assigned to SIV versus 64 (50%) assigned to placebo at visit one. No participants reported life-threatening (grade 4) events.

Among 142 BNT162b2 booster dose one and 43 BNT162b2 booster dose two recipients, the posterior median risk difference of grade ≥ 2 adverse events for SIV versus placebo recipients was 13% (95%CrI –0.03 to 0.27; probability of non-inferiority 62.7%) and -13% (95%CrI –0.37 to 0.12; probability of non-inferiority 98.5%). Among 18 mRNA1273 booster dose one and 35 mRNA1273 booster dose two recipients, the posterior median risk difference of grade ≥ 2 adverse events for SIV versus placebo recipients was 6% (95%CrI –0.29 to 0.41; probability of non-inferiority 69.2%) and –4%(95%CrI –0.30 to 0.23; probability of non-inferiority 91.4%) (table 2).

**Table 2.** Primary outcome: frequency of adverse events after coadministration of SIV or placebo with COVID-19 mRNA vaccines.

| Strata | Trial population | Total number of moderate or severe adverse events in intervention group | Total number of moderate or severe adverse events in placebo group | Posterior risk difference median (95%CrI) | Probability of non-inferiority |
| --- | --- | --- | --- | --- | --- |
| **BNT162b2**<br>**Booster dose one** | ITT | 43 (64%) | 35 (51%) | 0.13 (-0.03, 0.27) | 0.627 |
|  | PP |  |  | 0.12 (-0.03, 0.27) | 0.664 |
| **BNT162b2**<br>**Booster dose two** | ITT | 8 (36%) | 11 (55%) | -0.13 (-0.37, 0.12) | 0.985 |
|  | PP |  |  | -0.16 (-0.39, 0.09) | 0.991 |
| **mRNA1273**<br>**Booster dose one** | ITT | 5 (71%) | 7 (70%) | 0.06 (-0.29, 0.41) | 0.692 |
|  | PP |  |  | 0.02 (-0.37, 0.38) | 0.769 |
| **mRNA1273**<br>**Booster dose two** | ITT | 6 (40%) | 9 (47%) | -0.04 (-0.30, 0.23) | 0.914 |
|  | PP |  |  | -0.06 (-0.33, 0.21) | 0.941 |

Local adverse events were most frequently reported on day one across both groups. Grade ≥ 2 injection site pain was reported by 60 (42%), 12 (27%), 11 (61%), 11 (31%) of BNT162b2 first booster, BNT162b2 second booster, mRNA1273 first booster, and mRNA1273 second booster recipients, respectively; 85 (35%) were reported in the COVID-19 vaccine injection site, 8 (7%) at the SIV injection site, and one (1%) at the placebo injection site (table 3 & figure 2).

**Table 3.**
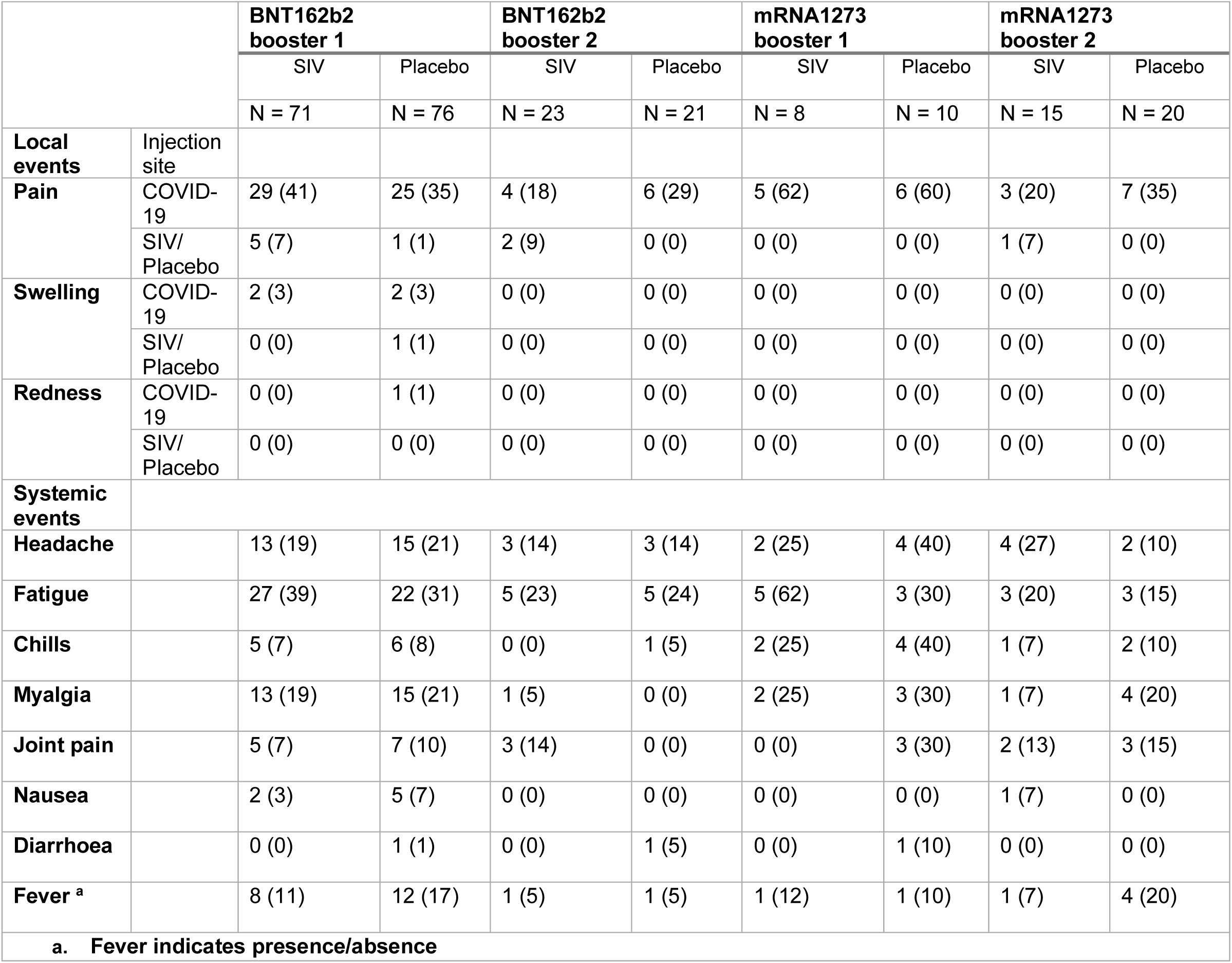
Frequency of moderate or severe local and systemic adverse events following visit one.

**Figure 2.**
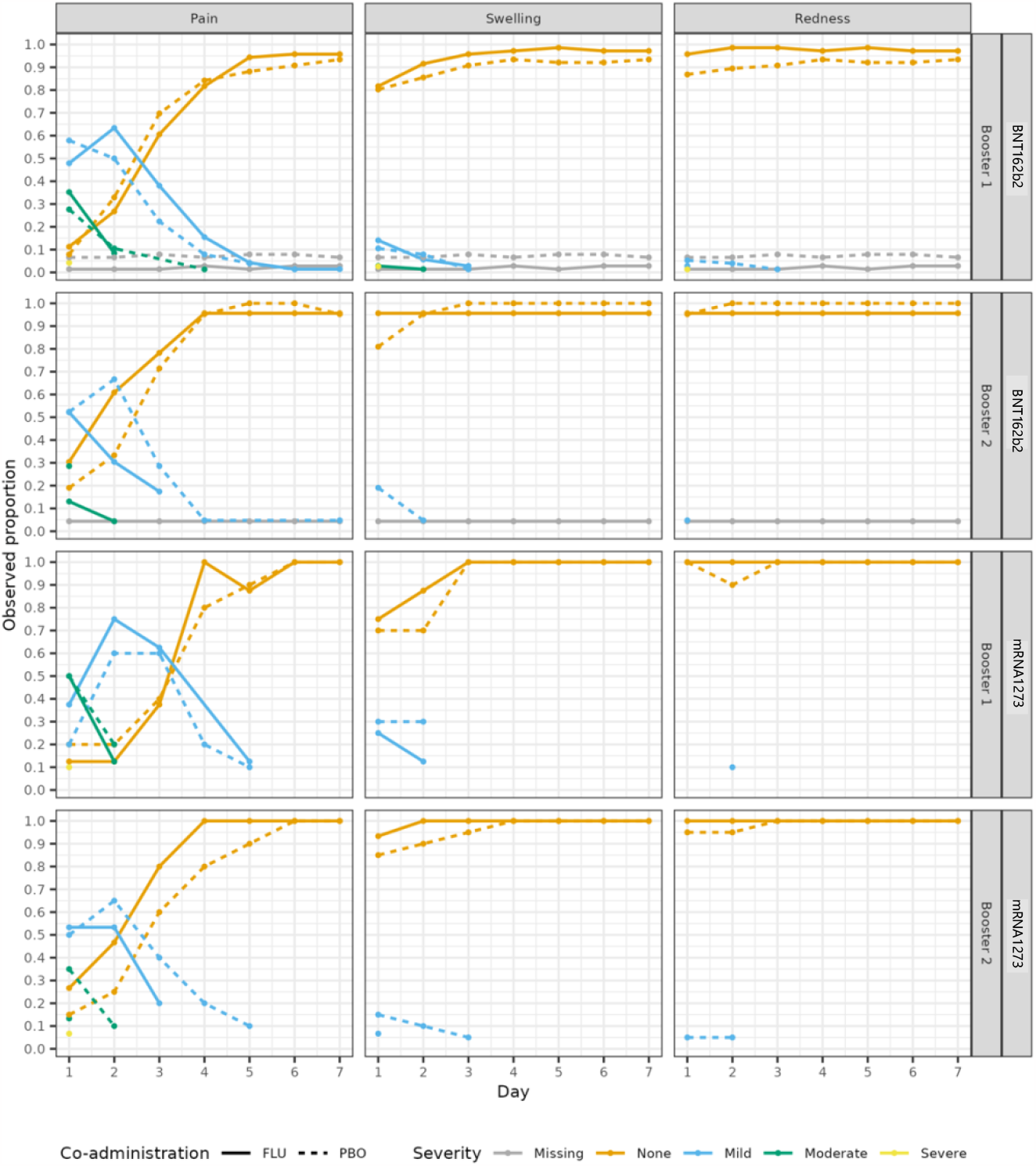
Observed proportion of participants co-administered SIV or placebo with local adverse events over 7 days by grade, COVID-19 vaccine type and dose number.

Systemic adverse events (where reported) were mostly reported as mild (table 3, figure 3 & 4). Of those reported, headache was the most common. In the placebo group, moderate or severe headache was reported by 15 (21%) first booster and by 3 (14%) second booster BNT162b2 recipients, and by 4 (40%) first booster and by 2 (10%) second booster mRNA1273 recipients. In the SIV group, moderate or severe headache was reported by 13 (19%) first booster and by 3 (14%) second booster BNT162b2 recipients, and by 2 (25%) first booster and by 4 (27%) second booster mRNA1273 recipients (table 3 & figure 3).

**Figure 3.**
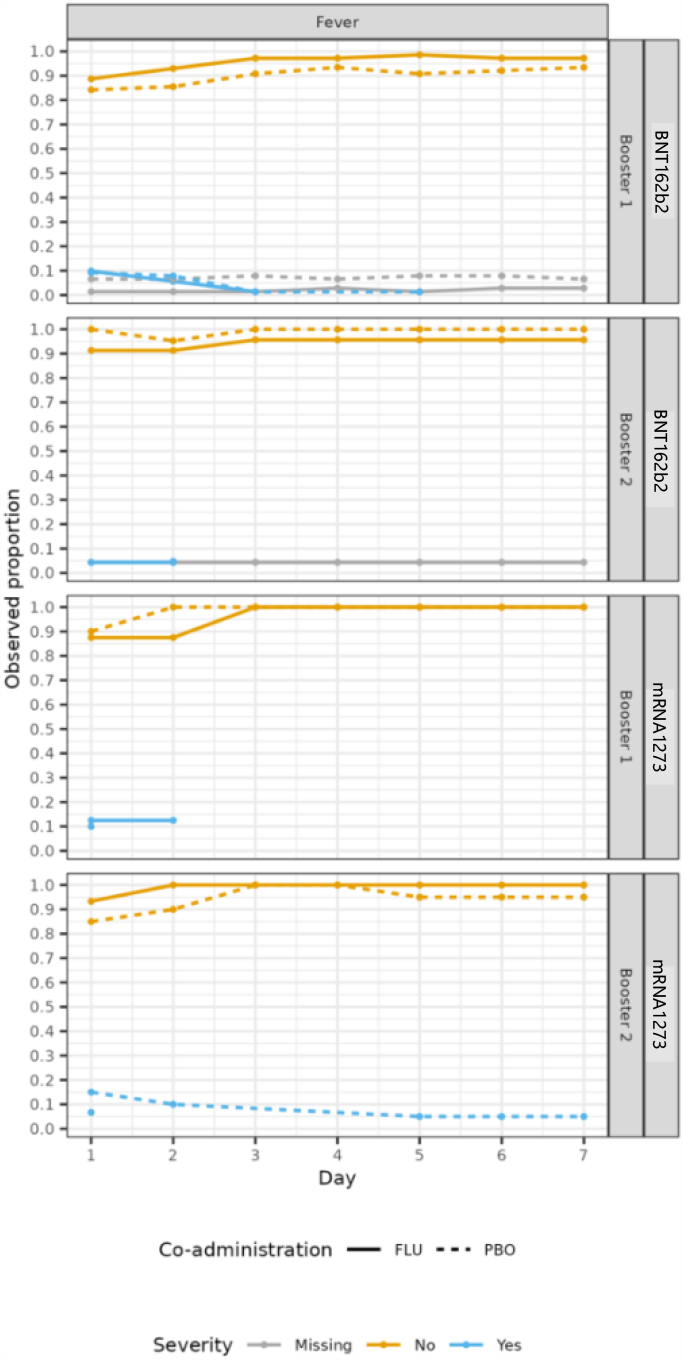
Observed proportion of participants co-administered SIV or placebo with fever over 7 days by grade, COVID-19 vaccine type and dose number.

**Figure 4.**
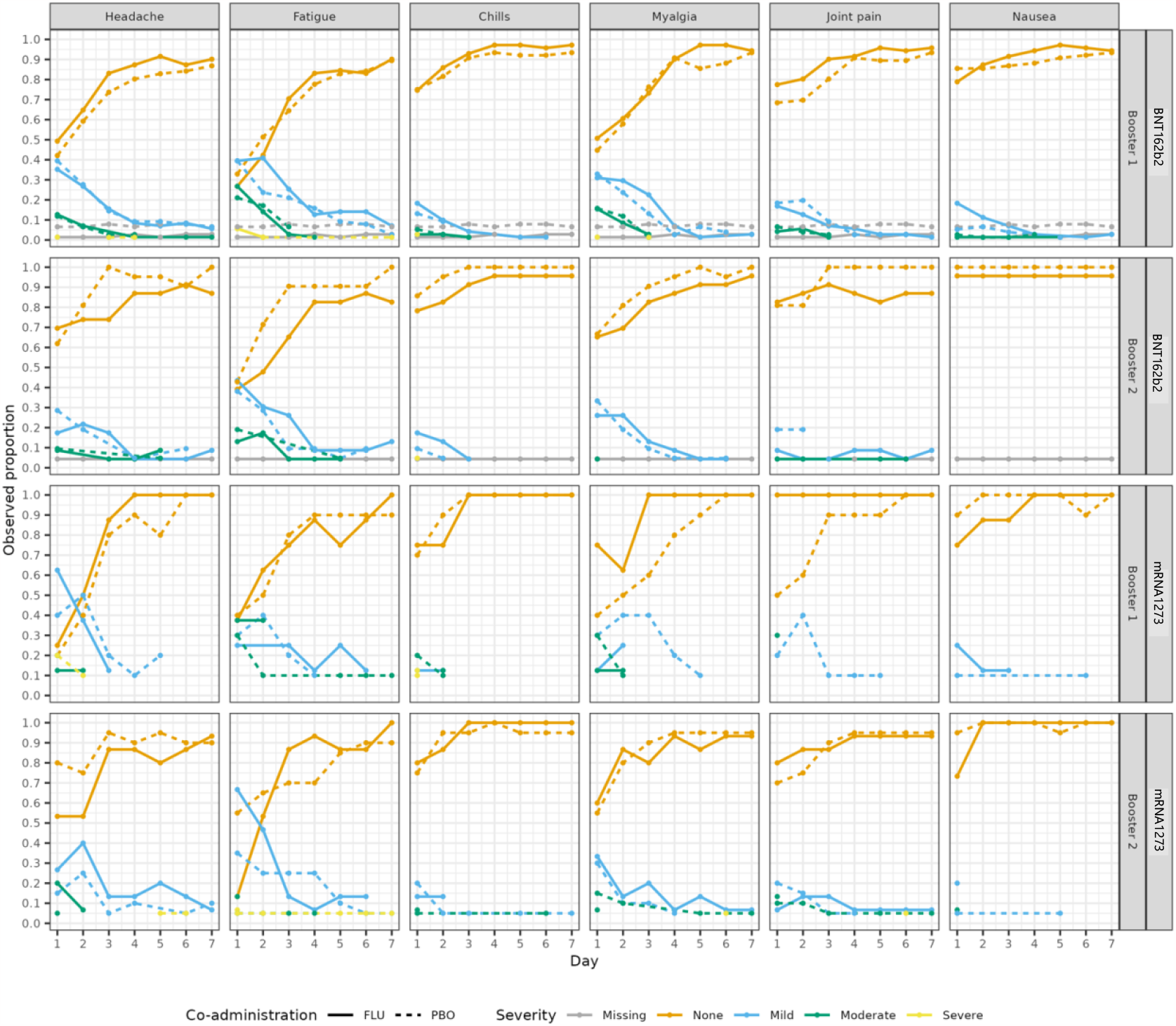
Observed proportion of participants co-administered SIV or placebo with systemic adverse events over 7 days by grade, COVID-19 vaccine type and dose number

One SAE, anaemia in a participant with a pre-existing condition, was ascertained in the 21 days post randomisation and was deemed unrelated to study vaccine (appendix F). No other medical attendances were ascertained within 21 days of enrolment. Two participants, one from either group, reported a COVID-19 infection within 21 days of enrolment; both were unable to adhere to study procedures owing to enforcement of infection control measures and therefore withdrew from the study.

Among 179 participants who were enrolled at baseline and who completed one or more surveys in the 7 days following enrolment, 37 (6%) who received SIV and 31 (5%) who received placebo at visit one reported one or more days absent from work.

## Interpretation and discussion

This randomised, placebo control trial investigated the safety and reactogenicity of COVID-19 and seasonal influenza vaccines. Existing published clinical trials evaluating the coadministration of SIVs with COVID-19 vaccines suggest the safety is acceptable (4,5,8,9). Local and systemic adverse events following coadministration are common and higher than observed when SIVs are administered alone (13,14), but similar in frequency to that observed following administration of COVID-19 vaccines alone (15). However, most trials to date explore non-mRNA vaccines or older age groups. Evidence informing the United States and Australian recommendations for the coadministration of COVID-19 vaccines and SIVs are based upon retrospective data. FluVID is a well-designed, blinded and controlled randomised trial evaluating the coadministration of two different mRNA vaccines with SIV or placebo. This trial significantly contributes to the pool of high-quality randomised trial data, enhancing our understanding of the safety of this approach.

We compared the frequency of moderate and severe adverse events following coadministration of SIV versus saline placebo among eligible and routinely vaccinated COVID-19 vaccine recipients. Most participants had received either their first or second booster doses of BNT162b2 or mRNA1273 immediately prior to visit 1. The pre-specified decision criterion for declaring non-inferior reactogenicity (<15% increased risk of moderate/severe adverse events) was only at the pre-specified decision threshold (>98.5%) for coadministration of SIV versus placebo in the context of a second booster dose of BNT162b2; the probability of non-inferiority of SIV versus placebo in the context of a second booster dose of mRNA1273 was 91.4% and did not meet our pre-specified criterion. We note that the small number of participants for other COVID-19 booster type/ dose number groups precluded meaningful evaluation of the pre-specified non-inferiority criterion.

The posterior median proportion of moderate/severe adverse events ranged from 0.38 (95% CrI 0.2 to 0.57) when SIV was co-administered with a second booster dose of BNT162b2, up to 0.77 (95%CrI 0.46 to 0.95) when SIV was co-administered with a first booster dose of mRNA1273. By comparison, the observed proportion of moderate/severe adverse events when SIV was administered alone (visit 2) was approximately 9%. This is comparable to Australian observational data of COVID-19 and SIV administered alone (16,17), suggesting that most of the reactogenicity was driven by COVID-19.

Our results for solicited adverse events are broadly consistent with other trials of the mRNA COVID-19 vaccines and SIV given alone (13,14,18,19), and in other trials which evaluated co-administration of these vaccines (4,8,9). In the ComFluCov trial, coadministration of a primary dose of BNT162b2 with an age-appropriate SIV or placebo in adults ≥ 18 years old, adverse events were of similar frequency, grade and duration to those observed in this trial (4). Similarly, in the TACTIC trial, which compared coadministration of a BNT162b2 booster with SIV to BNT162b2 booster and a placebo, SIV and a placebo, implied that there was no clinically relevant increase in adverse events when SIV and a BNT162b2 booster are co-administered (9). In another study that evaluated the coadministration of a mRNA1273 booster with a high dose quadrivalent influenza vaccine (QIV-HD) in adults ≥ 65 years old; the frequency of solicited adverse events in the mRNA1273 alone and coadministration group was similar, and higher than when QIV-HD was administered alone (8).

Our focus was on moderate or severe local and systemic adverse events, but the majority of the adverse events reported were mild (grade 1). Few severe (grade 3) adverse events and no life threatening (grade 4) adverse events were ascertained. Participants who received SIV at visit 1 were more likely to report mild pain and a mild headache compared to those who received a placebo. An increase in mild adverse events following co-administration of SIV with COVID-19 vaccines was reported in a study of 431 adults after coadministration with NVX-CoV2373 (5) and in a large cohort study of in the USA after coadministration with mRNA1273 or BNT162b2 (20).

This trial had several limitations. Enrolment stopped before the first planned analysis because of slow recruitment. We only enrolled participants who had received BNT162b2 or mRNA1273 vaccines; these were the predominant COVID-19 vaccines delivered in the Australian program at the time of enrolment and were the only vaccines routinely available at the community vaccination centres during the study period. Even so, few participants had received mRNA1273 vaccines. We monitored for serious adverse events and for work absenteeism, but these were too uncommon and enrolments too few to allow us to exclude an important difference between co-administered SIV versus placebo.

Despite these limitations, FluVID contributes to the literature regarding the reactogenicity caused by coadministration of SIV with COVID-19 vaccines. Our data are consistent with the moderate frequency of moderate and severe adverse events caused by BNT162b2 booster doses reported elsewhere (15). When SIV is co-administered, our data indicate a modest and acceptable increase in frequency of moderate and severe adverse events albeit only conclusive for second booster doses of BNT162b2. When SIV and COVID-19 vaccines are co-administered, vaccine recipients might incorrectly attribute adverse events to SIV and this might negatively affect future vaccine intentions. Vaccine providers should therefore counsel recipients of the risk of adverse events attributable to each vaccine alone and in combination to counter hesitancy toward future SIV doses. Nevertheless, on the balance of the safety and reactogenicity results from this and other studies, SIV and booster doses of the mRNA COVID-19 vaccines can be co-administered without concerns of a clinically meaningful increase in adverse events.

## Supporting information

Appendix A: FluVID study protocol V5.0

Appendix B: Daily diary card

Appendix C: Day 21 survey

Appendix D: Statistical analysis plan

## Data Availability

All data produced in the present study are available upon reasonable request to the authors.

## Ethics approval

The trial was approved by the Sydney Local Health District (Royal Prince Alfred Hospital) Ethics Committee (2021/ETH01232) and was sponsored by Sydney Local Health District.

## Acknowledgements

We would like to acknowledge the (Australian) National COVID-19 Consumer Reference Group for their review of the trial concept and design, James Totterdell for providing quality control of the analysis, the Data Safety and Monitoring Committee for their oversight of participant safety, the assistance of the Sydney Local Health District-Royal Prince Alfred Pharmacy and Vaccination teams, the staff at both the Mallett Street RPAH and Sydney Olympic Park Vaccination Centres, and the Sydney Local Health District-Royal Prince Alfred Ethics and Governance team.

JAR and MAJ are supported by the Snow Medical Research Foundation; MAJ was supported by NSW Health Office for Health and Medical Research; TLS is supported by an MRFF Investigator Award; PCMW is supported by an MRFF Investigator award.

We would like to thank the participants of the FluVID study for their engagement and participation in the trial.

## Author contributions

JAR and TLS was primarily responsible for drafting this manuscript. JAR, MAJ, WJB, TLS and IDC conceived the FluVID trial. MAJ and TLS designed the trial and MAJ was primarily responsible for the statistical analysis plan and performed the analysis. JAR, MAJ, AMDM, SLH, PCMW, MM, NW, KM, FJL, WJB, TLS and IDC have substantially contributed to the development of this project, and critically reviewed and approved the final version of this manuscript.

## Competing interests

All authors report no conflicts of interest.

**Appendix A:** FluVID study protocol V5.0

**Appendix B**: Daily diary card

**Appendix C**: Day 21 survey

**Appendix D:** Statistical analysis plan

