## Appendix A: FluVID study protocol V5.0 for "A single blinded, phase IV, adaptive randomised control trial to evaluate the safety of coadministration of seasonal influenza and COVID-19 vaccines (The FluVID study)"

**Study Title:** A single blinded, phase IV, adaptive randomised control trial to evaluate the safety of co-administration of seasonal in**flu**enza and CO**VID**-19 vaccines

**Short Title:** The FLU-VID study

**Coordinating Principal Investigator:** Professor Ian Caterson

**Sponsor:** Sydney Local Health District

**HREC Number:** 2021/ETH01232

**Protocol No:** X21-0209

**ANZCTR number:** 382319

***Open Source License****: this document was created under the Creative Commons Attribution Non-commercial ShareAlike Licence version 3.0 (http://creativecommons.org/licenses/by-nc-sa/3.0 ). It is freely available to be copied, adapted, distributed and transmitted under the conditions that: a) the original source is attributed; b) the work is not used for commercial purposes; c) any altered forms of this document are distributed freely under the same conditions.*

#### INVESTIGATOR SIGNATURE PAGE

The signature below constitutes the approval of this protocol and the attachments, and provides the necessary assurances that this study will be conducted according to all stipulations of the protocol, including all statements regarding confidentiality, and according to local legal and regulatory requirements and applicable Australian federal regulations and International Council for Harmonisation (ICH) guidelines.

**Coordinating Principal Investigator:**

Professor Ian D Caterson

Boden Professor of Human Nutrition, SoLES, University of Sydney
Medical Lead, RPA COVID Vaccination Clinic
Academic Clinical Director, CPC RPA Clinic
Deputy Clinical Stream Director, ACCR, SLHD

**Signed & Dated:**

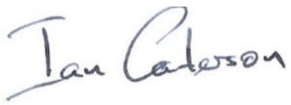

15/06/2021

### 1. Key Roles and Contact Information

| | |
|---|---|
| **Coordination Principal Investigator** | Professor Ian Caterson |
| **Sponsor** | Sydney Local Health District |
| **Associated Research Organisations** | Health and Clinical Analytics Lab<br>Sydney School of Public Health<br>University of Sydney<br><br>National Centre for Immunisation Research and Surveillance of Vaccine Preventable Diseases<br>Sydney Children's Hospital Network |
| **Study Sites** | Mallett Street Vaccination Centre<br>Royal Prince Alfred Hospital<br>94 Mallett St<br>Camperdown,<br>NSW 2050<br><br>NSW Health Vaccination Centre, Sydney Olympic Park<br>1 Figtree Drive<br>Sydney Olympic Park,<br>NSW 2127 |
| **Site Principal Investigators** | Site investigator: Professor Ian Caterson<br>Royal Prince Alfred Hospital Vaccination Centre<br><br>Site Investigator: Professor Ian Caterson<br>NSW Health Vaccination Centre, Sydney Olympic Park |
| **Trial Statistician** | Mark Jones |

| | |
|---|---|
| | University of Sydney<br> |
| **Study Investigators** | Professor Warwick Britton – Sydney Local Health District<br>Dr Frederick Lee – Sydney Local Health District<br>Professor Tom Snelling – University of Sydney<br>A/Professor Nicholas Wood – University of Sydney<br>Professor Kristine Macartney – University of Sydney<br>Mr Mitch Messer – Community representative<br>Dr Phoebe Williams – University of Sydney |
| **Funding** | Sydney Local Health District<br>Snow Medical Research Foundation |

A delegation of responsibilities and training log will be stored securely in the trial master file (TMF) in accordance with Good Clinical Practice (GCP).

#### 2. Standardisation

The protocol aligns with the standards set out by the Platform for evaluating Immunisation against COVID-19 (PICO) project and the PICO Core Protocol (1). This project aims to standardise the capture of data about, and biospecimens from, people undergoing immunisation against COVID-19. The data will be used to characterise the immune responses and clinical outcomes in an individual following immunisation, as well as identify any baseline factors which might influence these outcomes. It is hoped that collecting data and biospecimens in a standardised manner will enable sharing of data and/or biospecimens and aggregation of results, ensuring analyses are more complete, powerful, and timely.

#### 3. Abbreviations

| | |
|---|---|
| AE | Adverse Event |
| AIR | Australian Immunisation Register |
| ATAGI | Australian Technical Advisory Group on Immunisation |

| | |
|---|---|
| CPI | Coordinating Principal Investigator |
| COVID-19 | Coronavirus disease 2019 |
| GCP | Good Clinical Practice |
| HREC | Human Research Ethics Committee |
| iDSMC | Independent Data Safety & Monitoring Committee |
| MMR | Measles, Mumps and Rubella |
| mRNA | messenger Ribonucleic Acid |
| SAE | Serious Adverse Event |
| SARS-CoV-2 | Severe acute respiratory syndrome coronavirus 2 |
| SIV | Seasonal influenza vaccine |
| SOP | Standard operating procedure |
| TGA | Therapeutic Goods Administration |
| TTS | Thrombosis thrombocytopaenia syndrome |
| WHO | World Health Organisation |

#### 4. Summary

This study aims to evaluate, pragmatically and efficiently, the safety (and reactogenicity) of seasonal influenza vaccines co-administered with licenced COVID-19 vaccines in Australia. Simultaneous administration of vaccines is common practice within vaccine programs for children and adults worldwide. Co-administration of COVID-19 vaccines with seasonal influenza vaccines, if proven to be safe and effective, offers significant advantages for health systems planning (including cost, administrative practicalities, and effective use of resources) and for increasing the uptake of both vaccines. These benefits may be substantial given the likely need for booster doses of COVID-19 vaccines. It is envisioned that this study will be extended to assess the safety and immunogenicity of other potentially co-administered vaccines.

| | |
|---|---|
| Study design | Bayesian Adaptive, randomised, trial |
| Study duration | 18 months |
| Interventions | Participants will each receive a single dose of a licensed seasonal influenza vaccine (SIV) and a dose of a saline placebo vaccine 7-1 days apart. The SIV will either be a high dose adjuvanted (for those 65 years old or over only) or non-adjuvanted quadrivalent vaccine. |
| Timing | Day 0: Administration of SIV (group 1) or placebo (group 2) within 6 hours of a scheduled COVID-19 vaccine dose (either priming dose 1 or 2, or a booster dose)<br><br>Day 7 - 14: Administration of placebo (group 1) or SIV (group 2) |
| Study population | Eligible and scheduled to receive a COVID-19 vaccine on the day of enrolment (priming dose 1 or 2, or a booster dose), and also eligible to receive SIV |
| Inclusion criteria | To be eligible a person must:<br>1. Be aged ≥ 18 years old<br>2. Have received, or must be eligible, willing, and scheduled to receive, a COVID-19 vaccine on the day of enrolment<br>3. Be eligible and willing to receive a SIV in accordance with the Australian Immunisation Handbook<br>4. Provide informed consent |
| Exclusion criteria | A person is not eligible if they:<br>1. Are unwilling or unable to adhere to follow-up<br>2. Are contra-indicated to receive a SIV<br>3. Have previously received a COVID-19 vaccine not licenced in Australia |

| | |
|---|---|
| | 4. Immunocompromised individuals receiving supplementary primary COVID-19 vaccine doses due to high risk of primary vaccine failure<br>5. Received the SIV within the preceding 6 months |
| Strata and subgroups | Participants will be categorised as belonging to a stratum defined by their COVID-19 vaccine type (brand) and dose number they are to receive on the day of enrolment (e.g. Comirnaty first dose)<br>For each assigned brand of SIV, and in each stratum, we will compare those co-administered SIV versus placebo with their COVID-19 vaccine dose, and those administered SIV versus placebo 7-14 days after their COVID-19 vaccine dose. |
| Aims | 1. To evaluate the reactogenicity profile of SIV when co-administered (within 6 hours) with a COVID-19 vaccine, compared to placebo when co-administered with a COVID-19 vaccine.<br>2. To reduce deaths and suffering from COVID-19 disease through the collaborative generation and timely dissemination of high-quality evidence to inform COVID-19 immunisation strategy. |
| Primary objective | To determine if the risk of moderate or severe (grade 2 or above) adverse reactions occurring over the 7 days following co-administration of SIV with COVID-19 vaccines is no worse (non-inferiority margin equal to an absolute increase in risk of 15%) than the incidence after co-administration of saline placebo with COVID-19 vaccine |
| Primary endpoint | Solicited adverse reactions up to 7 days following co-administration of COVID-19 with influenza vaccine or placebo |
| Secondary objectives and endpoints | *1. Reactogenicity:* To compare the reactogenicity and safety of SIV co-administered with COVID-19 vaccine (within 6 hours) with the reactogenicity and safety of saline placebo co-administered with COVID-19 vaccine, and with SIV administered 7 to 14 days later<br>a. Solicited local reactions, assessed at days 0-7 after each visit<br>b. Solicited systemic reactions, assessed at days 0-7 after each visit<br>c. Serious adverse event(s) up to 21 days after visit 1 |

| | |
|---|---|
| | d. Medical attendance, up to 21 days after visit 1<br>e. Days off work (for participants in employment) up to 7 days after each visit<br>2. *Efficacy:* To determine whether the effectiveness of SIV and COVID-19 vaccine is no worse (non-inferiority margin equal to a relative increase in risk of 15%) when they are co-administered (within 6 hours) than when SIV is administered 7-14 days after COVID-19 vaccine.<br>a. Laboratory-confirmed COVID-19 infection up to 6 months after visit 1<br>b. Influenza-like illness or laboratory confirmed influenza infection up to 6 months after visit 1 |
| Number of participants | Maximum 1000 participants (the design allows for the sample size to be expanded if the need arises and additional resources become available) |
| Randomisation | Computer generated permuted block randomisation list by site and by COVID-19 vaccine type and dose number and SIV vaccine type with balanced allocation across treatment arms will be prepared by the trial statistician. |
| Blinding | Participants will be blinded to the group assignment. Research nurses administering the vaccines will be aware of the group assignment.<br>Research staff involved in the follow-up of adverse reactions or collecting specimens and laboratory staff processing specimens and performing immunogenicity assays will be blinded.<br>Researchers involved with the trial design modifications will be blinded to trial results unless a trial conclusion is reached.<br>Trial analysts preparing interim reports and the DSMC will be unblinded and unmasked. |
| Analysis | Generalised linear models fitted within a Bayesian framework |

### 5. Background

##### 5.1 COVID-19

Coronavirus disease 2019 (COVID-19) caused by the severe acute respiratory syndrome coronavirus 2 (SARS-CoV-2) has spread to populations around the globe, leading to significant numbers of hospitalisations and deaths (2). As of May 2021, the pandemic has seen over 150 million confirmed cases and caused over 3 million deaths (3). These numbers however, are likely to be a gross underestimation of the true burden of disease, with some reports predicting the global deaths to be more than double what has been reported (4). Indisputably, the worldwide spread of the virus has had devastating effects on populations, healthcare systems and economies.

While most of those infected with SARS-CoV-2 are asymptomatic or experience only mild illness, acute infection or COVID-19 can lead to intensive hospitalisation, long-term complications, or death (2,5–7). Such a wide spectrum of disease has created numerous challenges in mitigating viral spread and understanding clinical management. It is becoming increasingly evident that challenges due to COVID-19 will dominate global communities long into the future.

##### 5.2 COVID-19 vaccines

While non-pharmaceutical interventions have been successful in containing national outbreaks of COVID-19 in countries like Australia and New Zealand, these measures are not economically or socially sustainable in the long term. Many countries continue to struggle with rampant transmission, and it is only a matter of time until the global population resumes travel and other activities that increase the potential for viral transmission. When this occurs, COVID-19 will become resurgent even in areas where containment was successful. Vaccines are seen as the only intervention that could allow for a return to normal life without overwhelming healthcare systems and causing excessive deaths.

Several COVID-19 vaccines have been proven safe and effective and have been in use since late 2020. As of the start of May 2021, over a billion doses had been administered worldwide and over 2.5 million of those vaccine doses have been administered in Australia. As of October 2021, the Therapeutic Goods Administration (TGA) has approved three, non-live COVID-19 vaccines for use in Australia, which we will describe below.

Pfizer’s Comirnaty (BNT162b2) is a lipid nanoparticle-formulated, nucleoside-modified mRNA vaccine that encodes for the trimerised SARS-CoV-2 spike glycoprotein. mRNA vaccines use the pathogen’s genetic code as the vaccine; this then exploits the host cells to translate the code and then make the target spike protein. The protein then acts as an intracellular antigen to stimulate the immune response. The mRNA is then degraded within days. BNT162b2 encodes the SARS-CoV-2 full-length spike protein, which is modified by two proline mutations to lock it in the prefusion conformation and allow for it to more closely mimic the intact virus with which the elicited virus-neutralizing antibodies must interact.. The vaccine RNA is formulated in lipid nanoparticles (LNPs) for more efficient delivery into cells after intramuscular injection. Phase 3 clinical trials of Comirnaty demonstrated this vaccine to be safe and effective (8) and roll out of Comirnaty in

Australia, despite initial limited supply, has seen no major safety signals and less than 40% of vaccine recipients reporting any adverse reactions as per AusVaxSafety surveillance. The dose of Pfizer BioNTech COVID-19 vaccine is 30µg contained in 0.3ml of the diluted vaccine and ATAGI recommendation for the primary schedule is two doses, given 3 – 6 weeks apart. The vaccine comes in a multi-dose vial without preservative, each vial containing 6 doses in 0.45mL and requires dilution with 1.8mL of sterile 0.9% NaCl without preservative into each multi-dose vial.

AstraZeneca's COVID-19 vaccine (ChAdOx1 nCOV-19) is an adenovirus vector vaccine that uses replication-defective attenuated chimpanzee adenovirus vectors to carry SARS-CoV-2 spike protein DNA. As with the mRNA vaccine, once vaccinated the recipient cells produce spike proteins and mounts an immune response. The ChAdOx1 nCoV-19 SARS-CoV-2 spike (S) surface glycoprotein uses a leading tissue plasminogen activator (TPA) signal sequence. S is a type I, trimeric, transmembrane protein located at the surface of the viral envelope, giving rise to spike shaped protrusions from the virion. The S proteins subunits are responsible for cellular receptor ACE-2 binding via the receptor-binding domain and fusion of virus and cell membranes, thereby mediating the entry of SARS-CoV-2 into the target cells. The S protein has an essential role in virus entry and determines tissue and cell tropism, as well as host range.ChAdOx1 nCoV-19 expresses a codon-optimised coding sequence for Spike protein from the SARS-CoV-2 genome sequence accession MN908947. ChAd is a non-enveloped virus, and the glycoprotein antigen is not present in the vector, but is only expressed once the genetic code within the vector enters the target cells. The vector genes are also modified to render the virus replication incompetent, and to enhance immunogenicity (9). Once the vector is in the nucleus, mRNA encoding the spike protein is produced that then enters the cytoplasm. This then leads to translation of the target protein which act as an intracellular antigen.

Phase 3 trials demonstrated this vaccine to be safe and effect (10), however since implementation, the AstraZeneca vaccine has been shown to be associated with thrombosis thrombocytopaenia syndrome (TTS) 4-20 days after vaccination (11) which is very rare, occurring in approximately 4 per million people and may be fatal in approximately 1 in a million vaccinated. As of April 2021, in Australia, the AstraZeneca vaccine is no longer recommended for adults less than 50 years, yet the benefits still outweigh the risks in older age groups. The dose of AstraZeneca COVID-19 vaccine is 0.5mL and the recommended primary schedule is two doses, given a minimum of 12 weeks apart. The vaccine should be administered intramuscularly. The AstraZeneca vaccine is supplied in packs of 10 vials. Each vial contains 10 doses of vaccine, and is a colourless to slightly yellow, clear to slightly opaque liquid. Each dose is prepared by withdrawing 0.5 mL from a vial in a sterile 1 mL or equivalent syringe.

Moderna's COVID-19 Vaccine SPIKEVAX (elasomerna) was provisionally approved by the TGA on August 9, 2021. SPIKEVAX is a lipid nanoparticle encapsulated mRNA vaccine encoding for the full-length SARS-CoV-2 spike protein modified with 2 proline substitutions within the heptad repeat 1 domain (s-2P). Modifications allow for a prefusion conformation that stabilise the spike protein. After intra-muscular injection the mRNA of the spike protein is taken up and translated by cells. The spike protein is then produced and presented by cells to elicit an immune response. The dose of SPIKEVAX used is 0.5mL. The vaccine is administered intramuscularly. The ATAGI recommendation for the primary schedule is two doses, given 4 – 6 weeks apart. SPIKEVAX is supplied in multidose vials containing 10 doses of vaccine and is a white to off white suspension.

Additional vaccines are undergoing assessment by the TGA in Australia, including Novavax's adjuvanted recombinant spike protein-based vaccine and Johnson and Johnson's human adenovirus-vectored vaccine.

The Australian government has committed to the purchase of Novavax COVID-19 Vaccine (NVX-CoV2373) subject to TGA approval. Novavax COVID-19 vaccine is a nanoparticle-based protein vaccine containing the full-length SARS-CoV-2 spike protein and a Matrix-M1 adjuvant, which acts to boost the immune response. The dose of Novavax COVID-19 vaccine is 0.5mL. The vaccine should be administered intramuscularly. ATAGI recommendations on appropriate dosing are pending. Phase 3 trials administered 2 doses 21 days apart. Schedule and packaging will be specified within the TGA approved product information.

From October 27$^{th}$, 2021, ATAGI provided support a single booster dose for those who completed their primary COVID-19 vaccine course ≥6 months ago.

Many programmatic questions concerning COVID-19 vaccines will continue to face health officials into the future. These questions are diverse and include topics such as the interchangeability or mixing of vaccine types across the dosing schedule, the safety and acceptability of concurrent administration of scheduled vaccines, the duration of immune protection and the timing and need for booster doses, and optimal implementation strategies for most protecting the Australian population and way of life into the future.

##### *5.3 Influenza & Influenza vaccines*

Influenza causes serious illness and death worldwide. This usually seasonal viral infection is contagious and can be particularly dangerous in young children and the elderly. While the threat of influenza has somewhat diminished with implementation of COVID-19 non-pharmaceutical interventions (12), the threat of substantial disease from influenza is still a major public health concern. To have high rates of circulating influenza and COVID-19 could be catastrophic. While influenza's seasonality isn't completely understood, international travel is likely to play a major role in the virus's ability to travel from winter to winter around the globe. As international borders open and travel resumes, it is likely we will see it's return. Our best and most cost-effective intervention against influenza infection is still vaccination.

Seasonal influenza vaccination is recommended for people over the age of 6 months old in Australia and is free under the National Immunisation Program for high-risk groups. Seasonal influenza vaccines are required annually due to their short-lived protection, and the antigenic shift and drift resulting from mutations of antigenic components of the virus allowing viral escape of pre-existing immunity (13). The ideal timing for vaccination is before the June when the influenza season usually starts, with peak protection occurring within the first 3 – 4 months with notable waning occurring after 6 months. Recommendations for the SIV encourage vaccination throughout the year if influenza viruses are circulating, and a valid vaccine is available. Revaccination later in the same year is not routinely recommended but may benefit some individuals due to high risk of exposure or severe influenza (14). Recent studies have demonstrated revaccination can extend the period of protection against influenza and the frequency of reactions are similar between those receiving 1 or 2 doses (15,16).

Influenza vaccines in Australia are either split virion or subunit vaccines prepared from purified inactivated influenza virus containing antigenic fragments from two influenza A viral lineages and two influenza B viral lineages dependent on recommendations from global surveillance. Standard quadrivalent inactivated influenza vaccines are recommended for people < 65 years and adjuvanted quadrivalent inactivated influenza vaccines are recommended for people ≥ 65. The MF59 adjuvanted quadrivalent inactivated influenza vaccine (Fluad Quad) quantitatively and qualitatively enhances the immune response by attracting inflammatory cells to the injection site and thereby establishing an immunostimulatory environment for rapid uptake and comprehensive processing of the vaccine antigens (17). Live-attenuated influenza vaccines are not currently in use in Australia.

Under current recommendations, the Australian Immunisation Handbook advises that people can receive influenza vaccines at the same time as most other vaccines. Co-administration of the adjuvanted influenza vaccine with the adjuvanted subunit zoster vaccine is not recommended as there is uncertainty concerning the safety of simultaneous administration of two adjuvanted vaccines. As of October 27th, 2021, ATAGI recommends that it is acceptable to co-administer a COVID-19 booster vaccine dose with an influenza vaccine.

##### *5.4 Simultaneous administration of vaccines*

Simultaneous vaccine administration, the administration of more than one vaccine on the same clinic day, at different anatomic sites, is common practice in children and adults. Concurrently administering all vaccines, for which a recipient is eligible for, is highly desirable from a programmatic perspective, and has been shown to improve the likelihood that a person is age-appropriately vaccinated (18). A study of a measles vaccination demonstrated that one-third of infections during an outbreak could have been prevented if an MMR vaccine had been concurrently administered during a vaccination visit (19) (20).

Concurrent administration of vaccines is encouraged in global guidelines, with few exceptions(21–23). Studies show that co-administering the most frequently used live and inactivated vaccines produces similar immune responses and adverse reactions compared to administering vaccines separately (24–27).Co-administering live attenuated influenza vaccine with varicella and MMR vaccines as well as inactivated influenza vaccine with pneumococcal polysaccharide vaccine has been shown to produce no interference of immune responses (28,29). Evidence also exists co-administration of hepatitis B and yellow fever and measles and yellow fever vaccines is safe and immunogenic (30–32). Despite the lack of direct data on simultaneous administration of some vaccines such as oral typhoid, many vaccine guidelines recommend their coadministration (33).

According to the United States' Advisory Committee on Immunisation Practices (34), of all the registered vaccines and their pairwise combinations, there are only two exceptions to the recommendation that due vaccines should be administered simultaneously. Firstly, for persons with anatomic or functional asplenia and/or HIV, pneumococcal conjugate vaccine (PCV) should be administered first and then diphtheroid conjugated quadrivalent conjugate vaccine should be administered four weeks later. This recommendation is based on a single immunogenicity study that showed that when given together, antibody concentrations for 3 serotypes of pneumococcus (subtypes 4, 6B, and 18C) were robust, but lower than when the vaccines were given separately (35). The clinical significance of this is unknown, and NHMRC recommend that if the two vaccines are administered together (19), a repeat dose of neither vaccine is needed (36). Secondly, in patients recommended to receive both PCV and pneumococcal polysaccharide vaccine, PCV13 should be administered first because patients receiving PCV13, 8 weeks (children 6 - 18 years) or 1 year (adults > 18 years) before pneumococcal polysaccharide produce better immune responses than when given together (37–39). In the case of the latter combination, it is thought that the presence of polysaccharide in PPV23 may induce relative hypo-responsiveness to the pneumococcal capsular antigens; no influenza or COVID-19 vaccines contain polysaccharides. In summary, immunological interference between simultaneously administered vaccines is very much the exception rather than the rule, and there is no theoretical basis for expecting that COVID-19 and influenza vaccines should interfere with each other (40).

Comirnaty and COVID-19 Vaccine AstraZeneca are not live vaccines and share no common active ingredients with 2021 seasonal influenza vaccines. Therefore, there are no specific indicators to suggest co-administration of TGA approved seasonal influenza and COVID-19 vaccines is likely to be ineffective or harmful. There are no known studies demonstrating the reactogenicity and immunogenicity of simultaneous administration of Comirnaty or COVID-19 vaccine AstraZeneca with influenza vaccines. There is however, one known human trial which evaluated the safety and immunogenicity of co-administration of an investigational adenoviral vector vaccine against the respiratory syncytial virus and a standard dose of non-adjuvanted trivalent seasonal influenza in healthy adults ≥60 years. This trial demonstrated common but comparable injection site reactions and systemic reactogenicity with influenza vaccine co-administration versus vector vaccine alone (41).

In Australia, ATAGI now recommends that it is acceptable to co-administer COVID-19 and seasonal influenza vaccines, however evidence on co-administration is still limited (38, 39) . There is a lack of strong theoretical concerns against safety and efficacy. ATAGI also highlights within their advisory document that logistical issues may arise during high throughput vaccination of COVID-19 vaccine (42). Therefore, this study aims to pragmatically evaluate the co-administration of licenced COVID-19 and seasonal influenza vaccines within Australia. However, on May 27th 2021, the Chief Medical Officer of Australia announced that the 14 day window previously recommended between administration of COVID-19 vaccines and other vaccines could be waived for residents and staff in residential aged care. This recommendation came with assurance that the effectiveness of co-administration of influenza and COVID-19 vaccines will not be impacted.

COVID-19 vaccination is unlikely to be exclusive to a primary series and may require annual booster doses much like the influenza vaccine. Simultaneous administration has substantial benefits to health systems planning, economically favours health personnel time and may also reduce discomfort for vaccine recipients and thereby increase update of both vaccines (43).

### 6. Study Objectives and endpoints

| Primary Objective | |
|---|---|
| **Objective** | **Endpoint** |
| To determine if the risk of moderate or severe (grade 2 or above) adverse reactions occurring up to 7 days after co-administration of SIV with COVID-19 vaccines is no worse (non-inferiority margin equal to an absolute increase in risk of 15%) than the incidence after co-administration of saline placebo with COVID-19 vaccine | Any solicited adverse reaction of severity grade 2-4 occurring up to 7 days following administration of influenza vaccine or placebo, with a COVID-19 vaccine. |
| **Secondary Objective** | |
| **Objective** | **Endpoint** |
| Safety | |
| To compare the frequency of solicited adverse reactions, serious adverse events and adverse events of special interest after co-administration of SIV with COVID-19 vaccine (within 6 hours), with their frequency after saline placebo is co-administered with COVID-19 vaccine, and with SIV administered 7-14 days later. | Solicited local reactions, assessed at days 0-7 after visit 1 and visit 2 |
| | Solicited systemic reactions, assessed at days 0-7 after visit 1 and visit 2 |
| | Any serious adverse event up to 21 days after visit 1 |
| | Any adverse events of special interest up to 21 days after visit 1 |
| | Any medically attendance up to 21 days visit 1 |
| | Any days off work for participants in employment up to 21 days after visit 1 |
| Efficacy | |
| To determine whether the effectiveness of SIV and COVID-19 vaccine is no worse (non-inferiority margin equal to a relative increase in risk of 15%) when they are co-administered (within 6 hours) than when SIV is administered 7-14 days after COVID-19 vaccine. | Laboratory-confirmed COVID-19 infection up to 180 days after visit 1 |
| | Influenza-like illness or laboratory confirmed influenza infection up to 180 days visit 1 |

### 7. Study design

##### *7.1 Design*

The study design is a flexible multi-centre Bayesian sequential multi-arm placebo-controlled trial with adaptive sample size and stopping rules for both inferiority and non-inferiority. The design is intended to provide timely, robust inference on the safety profile a of co-administered COVID-19 with other vaccines and permits the introduction of new arms as COVID-19 vaccines become approved for use in Australia. The trial is configured to compare co-administration of SIV with COVID-19 (at prime/boost dose) versus co-administration of placebo with COVID-19 vaccine (at either dose). Figure 1 depicts a participant journey from randomisation through to exiting the trial.

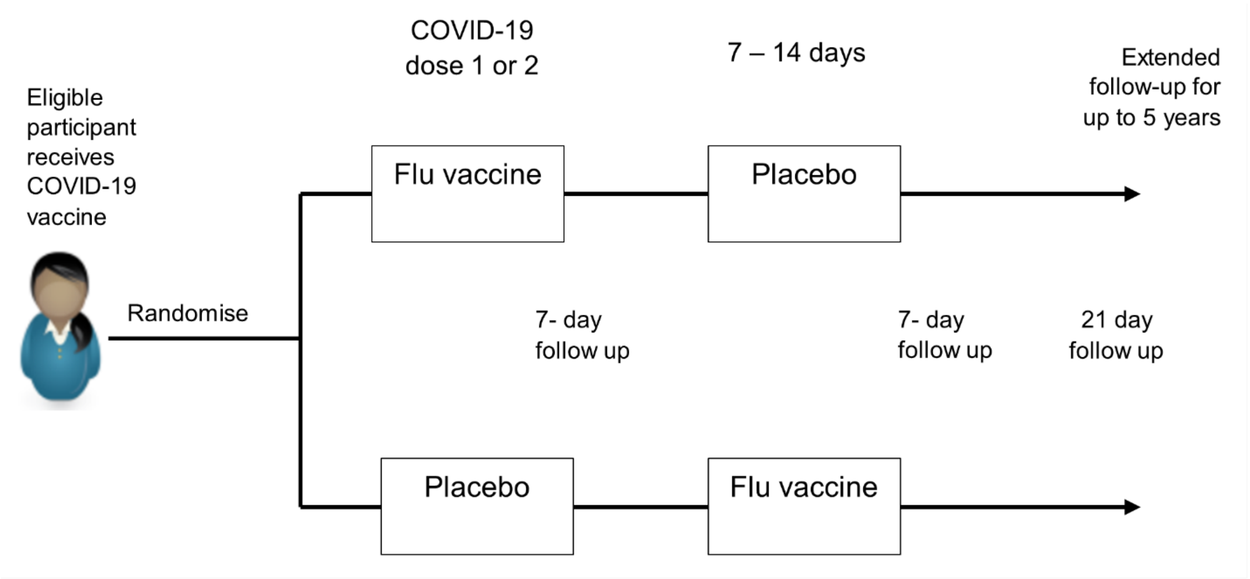

*Figure 1: Participant journey for evaluation of co-administered COVID-19 with influenza vaccine versus COVID-19 with placebo*

Participants who have received a first, second or booster dose of a COVID-19 vaccine are randomised to the intervention arms, initially being:

- SIV (within 6 hours of COVID-19 vaccine) followed by placebo 7-14 days later
- Placebo vaccine (within 6 hours of COVID-19 vaccine) followed by SIV 7-14 days later

Participants can only be involved in the trial once

##### *7.2 Expected participant numbers*

While the adaptive trial is designed to allow enrolment for as long as needed to reach a conclusion, current simulations have been made on the assumption that a maximum of 1000 participants will be enrolled across all COVID-19 brands and doses. Within the Australian setting, and the available resources, this is considered feasible and can be achieved without creating an undue burden on research staff.

As the trial employs stopping rules, the true sample size is a random variable. By simulation, the sample size has a median value of 700 participants across the simulated scenarios when considering two COVID-19 vaccine brands each having a two-dose schedule and assuming equal allocation to each. However, the median sample size falls to approximately 300 if only one brand is considered. Simulated scenarios ranged from all brand/dose pairs having a higher adverse event rate when co-administered with SIV to all brand/dose pairs having the same rate of adverse events. Specifically, in the scenario where all brand/dose pairs have a 30% increase in the adverse event rate when co-administered with a SIV, the cumulative probability of stopping the trial due to inferiority is above 90% by the time 600 participants are enrolled. In the null case, where there is no difference between the brand/dose pair adverse event rate irrespective of whether the COVID-19 vaccine is delivered with SIV or placebo, the cumulative probability of stopping is around 70% by the time that 900 participants are enrolled.

The simulations used a logistic regression model and assume decision thresholds of 0.985 for both inferiority and non-inferiority, interim analyses start when any single COVID-19 brand has 200 enrolled participants that have reached their 7-day follow up following visit 1. Subsequently, interim analyses are run for every additional 100 participants enrolled that have reached their 7-day follow up to any COVID-19 brand/dose pair up to a maximum of 1000 participants (in total across all brand/dose pairs). Additional assumptions include a baseline probability of adverse events associated with the COVID-19 vaccine were equal to 0.4, a minimum clinically important difference (non-inferiority margin) for moderate-severe reactions of 15% (in absolute terms) simulated differences between groups ranging from 0 (null case) to 30%, 1:1 enrolment across the first, second and booster dose of the COVID-19 vaccine, and even enrolment to COVID-19 vaccine brands. Given the dynamic nature of the COVID-19 situation, the trial simulation work is provisional and ongoing as information regarding the national vaccine roll-out of COVID-19 and seasonal influenza vaccine unfolds and new scenarios are identified. However, modifications to the design after the trial has commenced can only be proposed by study team members blinded to the accruing data and results.

##### *7.3 Time period of study*

Expected enrolment period for this study is 18 months.

##### *7.4 Study setting*

Multicentre study conducted through COVID-19 vaccination sites. Opportunistic enrolment at sites (including vaccination hubs) of COVID-19 vaccine administration. The initial sites are expected to be vaccination hubs operated by the Sydney Local Health District, namely Mallett Street Vaccination Centre, Royal Prince Alfred Hospital campus and NSW Health Vaccination Centre, Sydney Olympic Park.

### 8. Study participants

This is a pragmatic study and therefore aims to include a study population that is representative of most COVID-19 and SIV vaccine-eligible Australians, including those with risk factors for disease.

##### *8.1 Inclusion Criteria*

To be eligible a person must:

1. Be aged 18 years old or over
2. Have received within 6 hours, or must be eligible, willing and scheduled to receive, a COVID-19 vaccine on the day of enrolment
3. Be eligible and willing for SIV in accordance with the Australian Immunisation Handbook and national recommendations
4. Provide informed consent

##### *8.2 Exclusion criteria*

A participant will be excluded if any of the following criteria are met:

1. Unwilling or unable to adhere to follow-up (including electronic completion of 7-diary card)
2. Contra-indicated to receive SIV.
3. Previously received a COVID-19 vaccine not licenced in Australia
4. Immunocompromised individuals receiving supplementary primary COVID-19 vaccine doses due to high risk of primary vaccine failure
5. Receipt of SIV within the preceding 6 months

Contra-indications to SIV are detailed in the Australian Immunisation Handbook. At the time of commencement of the trial the contra-indications are:

- anaphylaxis after a previous dose of any influenza vaccine
- anaphylaxis after any component of an influenza vaccine

Precautions are described for individuals with a known egg allergy, latex allergy, history of Guillain-Barré syndrome and individuals receiving immuno-oncology therapy. The Handbook also advises deferring immunisation in people with acute systemic and febrile illnesses ≥ 38.5C).

##### *8.3 Strata and subgroups*

Participants will be categorised to one stratum based on the type of COVID-19 vaccine(s) and dose number received on the day of enrolment (e.g. Comirnaty first dose). Subgroups will be defined by the SIV type (brand) received..

For each assigned brand of SIV, and in each stratum, we will compare those co-administered the SIV with their COVID-19 vaccine dose, and those administered the SIV 7-1 days after their COVID-19 vaccine dose.

Over time, additional strata may be added, for example to accommodate new COVID-19 vaccines, booster (third or subsequent) doses, or heterologous COVID-19 vaccine schedules.

### 9. Treatment of study participants

#### *9.1 Description of study vaccines*

This is a Phase IV clinical trial and all vaccines used in this study are licensed for use by the Therapeutic Goods Administration in Australia.

##### *9.1.1 Seqirus Fluad Quad*

Fluad Quad is an adjuvanted quadrivalent inactivated influenza vaccine propagated in egg containing 60 µg of haemagglutinin per 0.5 mL dose. Each vaccine contains influenza surface antigens (haemagglutinin and neuraminidase) representing four influenza virus types as recommended by the WHO representing the viruses expected to be in circulation during the Southern Hemisphere winter. An antibody response to these surface antigens allows for protection against clinical disease in a high proportion of vaccine recipients. Fluad Quad is adjuvanted with squalene-based MF59 which allows for improved protection due to a greater overall immune response. The oil-in-water emulsion adjuvant enhances uptake and differentiation of antigen presenting cells against the vaccine antigens leading to qualitative and quantitative improvements in the immune response.

###### 9.1.1.1 Dosage, scheduling and packaging

One dose of Fluad Quad is 0.5mL. Prefered administration route is intramuscular injection. Vaccines should be shaken gently before administration. Fluad Quad comes in two packaging types; AUST R 313724: is a 0.5 mL suspension for injection in a needle-free pre-filled syringe (type I glass) or AUST R 316323: is a 0.5 mL suspension for injection in a pre-filled syringe (type I glass) with attached needle. Package sizes are 1’s or 10’s. Normal appearance of the vaccine is a milky-white suspension.

##### *9.1.2 GlaxoSmithKline Australia Fluarix Tetra*

Fluarix Tetra is a quadrivalent inactivated split virion influenza vaccine propagated in egg containing 60 µg of haemagglutinin per 0.5 mL dose. Each vaccine contains influenza surface antigens (haemagglutinin and neuraminidase) representing four influenza virus types as recommended by the WHO representing the viruses expected to be in circulation during the Southern Hemisphere winter. An antibody response to these surface antigens allows for protection against clinical disease in a high proportion of vaccine recipients.

###### 9.1.2.1 Dosage, scheduling and packaging

One dose of Fluarix Tetra is 0.5 mL. Vaccines should be given by intramuscular injection. Vaccines should be shaken before use and appear as a slightly opalescent suspension. Vaccines are supplied in a 0.5mL pre-filled PRTC syringe without needle. Package sizes are 1’s or 10’s.

##### *9.1.3 Saline placebo*

0.5 mL Sodium chloride 0.9% injection.
Study nurses will draw up 0.5mL saline syringes as required and label with a white opaque sticker.

##### *9.1.4 Sanofi Vaxigrip Tetra*

Vaxigrip Tetra is a quadrivalent inactivated split virion influenza vaccine propagated in egg containing 60 µg of haemagglutinin per 0.5 mL dose. Each vaccine contains influenza surface antigens (haemagglutinin and neuraminidase) representing four influenza virus types as recommended by the WHO representing the viruses expected to be in circulation during the Southern Hemisphere winter. An antibody response to these surface antigens allows for neutralisation of influenza viruses.

###### 9.1.4.1 Dosage, scheduling and packaging

One dose of Vaxigrip is 0.5mL. Vaccines should be given by intramuscular injection. Vaccines should be shaken before use and should appear as a colourless opalescent liquid. Vaccines are supplied in a pre-filled syringe with attached needle or with one separate needle or no needle provided per syringe. Package sizes are 1’s or 10’s.

##### *9.1.5 Sanofi Fluzone High-Dose Quadrivalent*

Fluzone High-Dose Quadrivalent is an inactivated split virion influenza virus vaccine propagated in egg containing 240 µg of haemagglutinin per 0.7 mL dose. Each vaccine contains influenza surface antigens (haemagglutinin and neuraminidase) representing four influenza virus types as recommended by the WHO representing the viruses expected to be in circulation during the Southern Hemisphere winter. An antibody response to these surface antigens allows for neutralisation of influenza viruses.

###### 9.1.5.1 Dosage, scheduling and packaging

One dose of Fluzone High-Dose Quadrivalent is 0.7mL. Vaccines should be given by intramuscular injection. Vaccines should be shaken before use and should appear as a clear and slightly opalescent liquid. Vaccines are supplied in a pre-filled syringe without attached needle. Package sizes are 5’s or 10’s.

##### *9.1.6 Sanofi FluQuadri*

FluQuadri is an inactivated quadrivalent split virion influenza vaccine propagated in egg containing 60 µg of haemagglutinin per 0.5 mL dose. Each vaccine contains influenza surface antigens (haemagglutinin and neuraminidase) representing four influenza virus types as recommended by the WHO representing the viruses expected to be in circulation during the Southern Hemisphere winter. An antibody response to these surface antigens allows for neutralisation of influenza viruses.

###### 9.1.6.1 Dosage, scheduling and packaging

One dose of FluQuadri is 0.5 mL. Vaccines should be given by intramuscular injection. Before administering a dose of vaccines, the prefilled syringe should be shaken. FluQuadri suspension for injection is clear and slightly opalescent in colour. Prefilled syringe (clear syringe plunger rod), 0.5 mL with or without separate needle. Package sizes are 5's or 10's.

##### *9.2 Storage of study vaccine and placebo*

The Investigator at each participating site is responsible for ensuring the maintenance and viability of the vaccines and their appropriate storage according to local procedures.

Influenza vaccines and placebos will be stored in small quantities in separate vaccine approved fridges. The fridge will be monitored, and procedures will be specified (e.g. batching of vaccines) within standard operating procedures to ensure the temperature fluctuations will be minimised.

##### *9.3 Administration of vaccines*

Standard vaccination practices will be observed. Administration must be performed within the clinics by an appropriately qualified, experienced and delegated study registered doctor or nurse from the study team between the operating days and hours of the trial (Monday to Friday, 8am to 4pm). To ensure that participants receive the correct dose of vaccines, the unblinded study staff must confirm and record the dose volume, batch number and date of administration in the relevant source documentation prior to administering the vaccine. The date and batch numbers of all vaccines administered will be recorded in the participant's personal immunisation record and the Australian Immunisation Register (AIR). Used syringes are to be disposed of appropriately.

Appropriate medication and other supportive measures for management of an acute hypersensitivity reaction should be available as per standard immunisation practices. Participants will be observed closely for 15 minutes following the administration of vaccines, with appropriate medical treatment readily available in case of a rare anaphylactic reaction.

##### *9.4 Compliance with Trial Treatment*

The study nurse or doctor who delivers the study vaccines or placebo will record if a dose is refused after randomisation has occurred.

##### *9.5 Accountability of Trial Treatment*

Unused vaccines, placebos and returned used vials will be discarded in accordance with local pharmacy protocols. The vaccines will be collected from the vaccine fridge at the beginning of each day scheduled for study visits. All used vaccine boxes will be returned to the study site at the end of each study day. The cold chain will be maintained, for example using ice packs in a cold box in order to ensure the temperature of vaccines is maintained at 2 - 8°C.

##### *9.6 Concomitant medication*

Participants will be advised that they may take paracetamol prophylactically after vaccine administration. This will be from the participants own supplies rather than supplied by the study team.

##### *9.7 Post-trial Treatment*

No further treatments will be provided by the sponsor to study participants after participant completion of the study period and exit from the trial.

### 10. Study procedures

##### *10.1 Study flow chart*

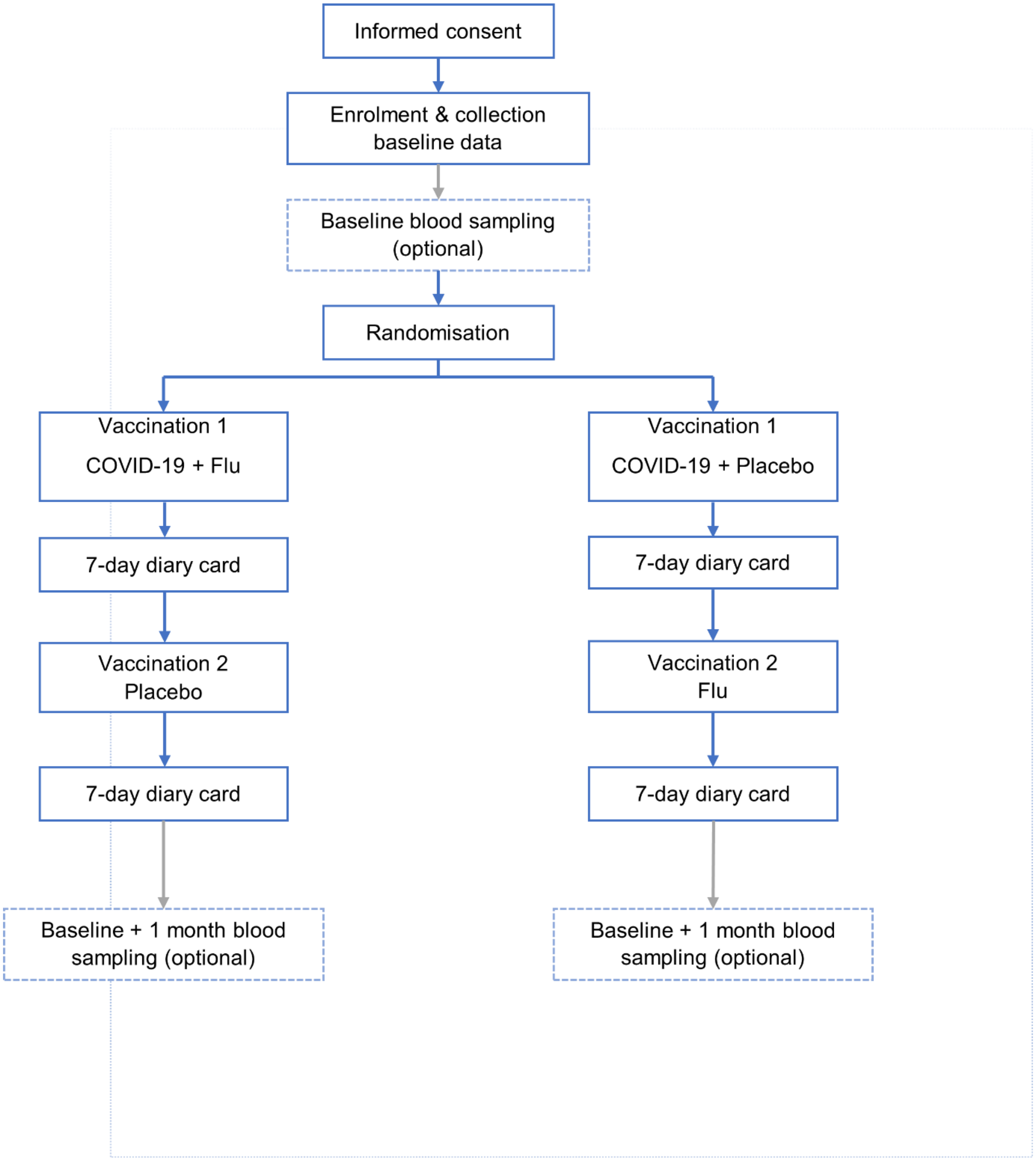

*Figure 2: Summary of study procedures and study flow*

##### 10.2 Identification of potential participants

Potential screening avenues to identify eligible participants include:

- *During immunisation clinic visits* - potential participants may be directly approached in the course of scheduled visits for COVID-19 immunisation at participating sites. Trained research staff will approach participants to discuss participation.
- *Advertising material* - including flyers and posters, may be used to promote and raise awareness; these may appear on public notice boards or electronic notice boards. Advertisements may also be displayed in electronic or print media and various community locations. Subject to HREC and governance approvals.
- *Text/email message* - potential participants may be provided with a link, within their vaccine appointment reminder, to the study advertising material and be invited to register their interest within the study REDCap database. Subject to HREC and governance approvals.

Research staff at participating sites will be trained on study requirements to ensure eligible participants are identified, educated about, and invited to participate in the study. Site agreements will stipulate that enrolment and screening procedures occur according to the approved protocol and in line with the principles of Good Clinical Practice (GCP) and Declaration of Helsinki.

Each site will maintain a screening log to capture basic demographic data on all individuals screened, eligible and enrolled, and reasons for non-eligibility. The purpose of the log is to allow full characterisation and representativeness of the study population and to identify any deficiencies in process.

##### 10.3 Informed consent

Informed consent will be obtained before data collection and other study related procedures begin. Electronic and/or written versions of the Participant Information Sheet and Informed Consent (PISIC) will be provided to the participant detailing no less than: the exact nature of the study; what is involved for the participant; the implications and constraints of the protocol; any risks associated with taking part; that anonymised samples may be shared with study collaborators; that individual results will not be shared with participants. It will be clearly stated that the individual is under no obligation to participate and is free to withdraw at any time, for any reason, and without prejudice to future care, and with no obligation to explain the reason for withdrawal.

Participants will be allowed a reasonable time period between the provision of information to prospective participants and the use of their data so that an opportunity for them to decline to participate is provided before the research begins. In order to maximise the opportunity for participants to consider their participation, where possible, they will be contacted at least 24 hours prior to their scheduled COVID-19 vaccination via SMS (wording now submitted with this application) or be informed of the study via advertisements and invited to register their interest in participation (the Flu-VID registration form has now been uploaded with this resubmission and will be collected in a REDCap project separate to the main study file). Those who indicate their interest will be provided the information contained within the participant information sheet.

Signage (submitted with this re-submission) at the vaccination centre will also be utilised to invite registration of interest on the day of COVID-19 vaccination by connection to the web-based study information and registration form. Those who connect will be provided with the participant information sheet in electronic form. A research nurse will discuss the information sheet and answer questions. Where possible, this will occur while waiting for COVID-19 vaccination; if not, it will occur immediately afterward in the 15 minute post-vaccination observation period.

While the risks of participating in this study are small compared to the types of decisions patients make every day in the context of a single clinic visit, we will aim to give potential participants as much time as possible to consider their enrolment in the study. However, there will necessarily be a window after COVID-19 vaccination to enrol and vaccinate the participant. While arbitrary, we believe that 6 hours represents a maximum interval in which two vaccines could be reasonably considered to be co-administered, practically representing administration on the same working day. Six hours therefore represents a maximum interval for a person to consent to participate after COVID-19 vaccination. Importantly, we will not enrol anyone who is unable to provide valid consent in this time. Participants who are attending for their first COVID-19 vaccine will be able to defer enrolment to their visit for their second COVID-19 vaccine.

Research staff will ensure that within this time a mutual understanding between researchers and participants is obtained and opportunities for participants to ask questions and to discuss the information and their decision with others, if they wish, are provided. Informed consent will be obtained by means of written or electronic signature which will be dated and countersigned by the person who obtained it.

The collection of blood samples is optional (see section 11.1) and will require opt-in consent. Participants will be explained the nature of the blood sampling and the additional risks and benefits associated with collection of blood samples to allow an informed choice.

This study will adhere to the principles of dynamic consent, whereby consent may be reassessed across the duration of the study to ensure it is accurate and up to date. Consent for any data and/or biospecimens collected to be used for future research purposes may also be invoked or withdrawn by the participant or legal representative over time.

##### *10.4 Screening and Eligibility Assessment*

Screening and eligibility will occur by a suitably qualified and delegated member of the research team. Inclusion and exclusion criteria (specified in section 8) will be checked following prior to randomisation and after the participant has provided informed consent.

##### *10.5 Randomisation, blinding and code-breaking*

###### *10.5.1 Randomisation*

Computer generated permuted block randomisation list stratified by site, by COVID-19 vaccine type and dose number, and by SIV vaccine type, with balanced 1:1 allocation across treatment groups will be prepared by the trial statistician.
On the day of scheduled COVID-19 vaccination, participants for whom all eligibility criteria have been satisfied, for whom informed consent for participation has been obtained and a unique participant number (generated from the REDCap Database) will be randomised once. A qualified and delegated member of the research team will record the participant's identifiers and a sequential participant number and then an unblinded research nurse will obtain the next contiguous allocation (i.e. the lowest available randomisation number) the study REDCap database. The randomisation allocation will be recorded in the CRF and at the point of randomisation the participant will be considered enrolled into the study.

###### *10.5.2 Blinding*

At each vaccination visit, an unblinded member of the research team will dispense and administer the study vaccine (SIV or saline placebo) to a blinded participant. Pre-filled syringes containing either the SIV or a saline placebo will be covered with opaque tape and concealed in an opaque box until ready for administration. Prior to opening the box the participant will be asked to look away. The member of the research team will record the assigned study number and administer the study vaccine intramuscularly per Australian guidelines.

Research staff involved in the follow-up of adverse reactions will be blinded to the group assignment of the participants.

Laboratory staff processing or analysing trial specimens will be blinded to the group assignment of participants.

Trial analysts preparing interim reports will be unblinded and unmasked.

###### *10.5.3 Code-breaking*

Randomisation code will be held by the trial statistician and will be password protected. The code will only be used if a situation arises where it is deemed necessary by the CPI to break the blinding process for compelling medical or safety reasons. The CPI may delegate this responsibility to another study investigator in his/her absence. The risk posed in the study is deemed to be small, and knowledge of the identity of the assigned treatment very unlikely to affect immediate management; therefore any code breaking will be conducted in normal business hours. Authorisation for unblinding will be given to the trial statistician by the CPI or the CPI's delegate. The statistician will then communicate directly with the medical team caring for the participant in order to maintain blinding of the study team. Participants for whom the assignment is revealed will remain in the study and where possible continue to receive any scheduled assigned vaccines unless medically contraindicated.

Where requested by a participant or by a clinician, unblinding of any participant may occur after the study database is formally locked for the final analysis. Should the study terminate early due to a decision by the CPI or the Sponsor, unblinding may be undertaken prior to completion of the database lock where necessary.

The Data and Safety Monitoring Committee (DSMC) may request of the CPI access to unblinding data at any point in the study.

##### *10.6 Visit descriptions*

Visit 1

Study Day 0: Enrolment and first vaccination

All participants

- Informed consent is obtained and documented either electronically or on paper
- Eligibility criteria will be checked prior to enrolment
- Baseline data collected
  - Demographic: date of birth, sex, occupation, ethnicity/Indigenous status
  - Medical History: height, weight, existing medical conditions (allergies, diabetes, cardiovascular disease, respiratory disease, immunocompromising disorder, liver disease, renal disease, cancer/malignant neoplasm, history of smoking, pregnant, concomitant medications, history of blood products, receipt of vaccines in the 30 days prior to enrolment, COVID-19 vaccine history including vaccine type, dose number, date of vaccination and, if known, batch number.
- Randomisation will be performed by an unblinded study nurse
- Vaccination:
  - Intramuscular administration of SIV or placebo vaccine, in the opposite arm to the previously administered COVID-19 vaccine
- Participants will be observed closely for at least 15 minutes, with appropriate medical treatment readily available in case of a rare anaphylactic reaction following the administration of vaccines
- Confirm participant phone number and explanation of visit 1 vaccine diary card to assess local and systemic adverse events for 7 days following vaccination sent daily via email/text. Participants will be provided with a tape measure, thermometer and verbal instructions for use
- Book appointment for visit 2

Optional blood sampling (Mallett Street Vaccination Centre only)

- For participants who opt-in to additional blood sampling, up to 20mLs of blood will be collected into a serum (clotted) tube (2 x 10mL) at baseline for future research (excluding genetic studies). Baseline bloods will be considered if collected no more than 72 hours prior and 6 hours after vaccination with SIV.

Visit 1 follow-up

- An electronic vaccine diary card will be sent to the participant daily from Day 1 to Day 7 following visit 1 to ascertain local and systemic adverse events and their grade.

Visit 2

Study Day 7 - 14

- Reporting of any SAEs that might have occurred since the last visit
- Review of any incomplete vaccine diary card entries
- Review medical history and check of contraindications
- Pre-vaccination assessment: Vaccination should be deferred in individuals with acute systemic and febrile illnesses ≥ 38.5C, and if possible re-scheduled for administration before the end of Day 14.
- Vaccination: intramuscular administration of SIV or placebo
- Participants will be observed closely for at least 15 minutes, with appropriate medical treatment readily available in case of a rare anaphylactic reaction following the administration of vaccines
- Explain visit 2 vaccine diary card to assess local and systemic adverse events for 7 days following vaccination sent daily via email/text.

*Note: a 7 - 14 day window has been selected to allow for flexibility of vaccine administration for those receiving Comirnaty vaccine dose 1. Guidelines currently recommend a 21 day window between dose 1 and dose 2, therefore those receiving dose 1 of Comirnaty should attend study visit 2 at least 7 days after dose 1 and 7 days before dose 2.*

Visit 2 follow-up

- An electronic vaccine diary card will be sent to the participant daily for 7 days following Visit 2 to ascertain local and systemic adverse events. The first entry will occur approximately 24hr after vaccination.
- Study nurse will contact participant approximately 7-14 days following visit 2 to review any incomplete diary card entries (if apparent) and obtain any SAEs that may have occurred since enrolment

Optional Visit 3

Study day 21 - 42

- For participants who opt-in to additional blood sampling, up to 20mLs of blood will be collected into a serum (clotted) tube (2 x 10mL) for future research (excluding genetic studies) at least 21 days after COVID-19 immunisation

21 day follow-up

- Participants will be sent an electronic survey (and followed-up with a phone call if required) on Day 21 to obtain any event that could be considered any AE that is serious (SAE), of special interest (AESI), change in medication (including any additional vaccines) or medical attendance.

##### *10.7 Recording of Vaccinations of AIR*

All immunisations administered to participants over the duration of the study will be recorded on the Australian Immunisation Record (AIR). Influenza immunisation records will be uploaded at the end of the participant's study period (after Day 21 follow is complete) by the unblinded vaccinator and will include the accurate date of vaccination.

##### *10.8 Discontinuation/Withdrawal of Participants from the Study*

A participant will have the right to withdraw from the study at any time and for any reason, and will be asked but not be obliged to give a reason for doing so.
Participants may be discontinued from the study if:

- The participant or legal representative request withdrawal from ongoing participation
- The Site Principal Investigator considers that continued participation is not in the best interests of the participant

If a participant withdraws from the study, storage of samples will continue unless the participant requests otherwise. Any data collected before their withdrawal will still be used in the analysis for safety and trial integrity. A participant may also be voluntarily withdrawn from the study due to what they perceive as an intolerable AE. If this occurs, the participant must undergo an end of study assessment and be given appropriate care under medical supervision until symptoms cease or the condition becomes stable. Withdrawn participants will be returned to the care of their general practitioner for the completion of any outstanding vaccinations.

##### *10.9 Follow up of withdrawn participants*

If a participant is withdrawn due to an adverse event, the Investigator will arrange for follow-up visits or telephone calls until the AE has resolved or stabilised. Attempts will be made to contact a participant who does not return for scheduled study visits.
Contact may be made via phone, text, email or mail. Information will be gathered about any solicited and unsolicited AE's that have occurred within 7 days of vaccination and any SAEs or AESIs that have occurred at any time during the study.
If the participant withdraws, long-term safety data collection, including some procedures such as safety bloods, may still continue as appropriate if they have received one or more vaccine doses, unless they decline any further follow-up.

Withdrawn participants will be informed of their allocated vaccination group 7 days after their last vaccine visit.

##### *10.9 Definition of End of Trial*

The end of the trial and trigger for reporting/publication of results is the date of completion of the 21 day follow up for the last enrolled participant. Long-term follow up and future research will be reported, however this will not restrict the release of the information pertaining to the primary endpoint.

### 11. Laboratory Methods

##### *11.1 Optional blood sampling*

Participants at Mallett Street Vaccination Centre will be invited to opt-in to additional blood sampling future research (excluding genetic studies) relating to the coadministration of COVID-19 vaccines with SIV or placebo. Participation in blood sampling is not required for participation in the trial. Participants will be required to provide additional consent.

*11.1.1 Blood sample*

Up to 20mLs of blood will be collected into a serum (clotted) tube (2 x 10mL) at the study site by qualified and delegated study staff.

*11.1.2 Timing*

Blood sample 1: Visit 1 with a window of 72 hours prior to vaccination and no more than 6 hours after vaccination with SIV

Blood sample 2: 21 - 42 days after Visit 1. The blood sample should be collected prior to any further doses of COVID-19 vaccine.

##### *11.2 Anonymisation of laboratory samples*

Samples will not be labelled with information that directly identifies the subjects but will be coded with the unique participant number for the subject. Labelled specimens will then be carried by study staff to The Centenary Institute Medical Research Foundation Laboratory for storage.

##### *11.3 Laboratory Processing and analysis*

*11.2.1 Collaborating laboratories*

The collaborating laboratories involved in this study include:

The Centenary Institute Medical Research Foundation
Building 93,
Royal Prince Alfred Hospital
Missenden Rd
Camperdown NSW 2050 Australia

*11.3.2 Specimen Sampling, Storage Procedures and Transport*

Study and site-specific operating procedures will be used for the collection, handling and storage of specimens. Any transfer of samples will occur under a Material Transfer Agreement and anonymised with unique coded identifiers to protect the identity of the participant. The transport conditions will be compliant with all regulations and guidance listed in the Australian Dangerous Goods Code Edition 7.6 (https://www.ntc.gov.au/sites/default/files/assets/files/Australian-Code-for-the-Transport-of-Dangerous-Goods-by-Road%26Rail-7.6.pdf).

##### *11.4 Future research & laboratory investigations*

Specific permission will be sought from participants to retain unused samples following completion of this study. Stored samples will be anonymised prior to storage. These samples will be retained with the intention of use in future research studies. Participants can refuse permission for these samples to be stored and, even if permission is granted initially, will be able to request that these samples be destroyed at any time.
The CPI will have custodial responsibility for these samples, and access to these samples will be at their discretion. Use of these samples will be pending ethical approval for use in another related project.

#### 12. Safety Reporting

##### *12.1 Definitions*

| **Term** | **Definition** |
|---|---|
| Adverse Event (AE) | Any untoward medical occurrence in a patient or clinical trial participant administered a medicinal product and that does not necessarily have a causal relationship with this treatment. |
| Solicited/Expected Adverse Event (S/EAE) | Solicited AEs are those events specifically captured in the 7 day diary card issues after each vaccination.<br><br>Solicited AEs collected for this study include: pain, induration, erythema, temperature, headache, fatigue, chills, myalgia, joint pain, nausea, diarrhoea. Solicited AE's (local and systemic) will be captured for a period of 7 days post-vaccination |
| Adverse Reaction (AR) | Any untoward and unintended response to an investigational medicinal product related to any dose administered.<br><br>**Comment***:* All adverse events judged by either the reporting investigator or the sponsor as having a **reasonable possibility of a causal relationship** to an investigational medicinal product would qualify as adverse reactions. The expression 'reasonable causal relationship' means to convey, in general, that there is evidence or argument to suggest a causal relationship. |

| | |
|---|---|
| Serious Adverse Event (SAE) | Any adverse event that results in death, is life-threatening, requires unplanned hospitalisation or prolongation of existing hospitalisation, results in persistent or significant disability or incapacity, or is a congenital anomaly or birth defect.<br><br>**Note:** Life-threatening in the definition of a serious adverse event or serious adverse reaction refers to an event in which the participant was at risk of death at the time of the event. It does not refer to an event that hypothetically might have caused death if it were more severe.<br><br>**Note:** Medical and scientific judgement should be exercised in deciding whether an adverse event/ reaction should be classified as serious in other situations. Important medical events that are not immediately life-threatening or do not result in death or hospitalisation, but may jeopardise the participant or may require intervention to prevent one of the other outcomes listed in the definition above should also be considered serious. |
| Serious Adverse Reaction<br>(SAR) | An adverse event that is both serious and, in the opinion of the reporting Investigator, believed with reasonable probability to be due to one of the trial treatments, based on the information provided. |
| Suspected Unexpected Serious Adverse Reaction (SUSAR) | An adverse reaction that is both serious and unexpected. |
| Adverse Events of Special Interest (AESI) | Adverse event identified as being of particular relevance to the study treatment. These will also be reported as an SAE if meeting the SAE criteria. |

*NB: to avoid confusion or misunderstanding of the difference between the terms "serious" and "severe", the following note of clarification is provided: "Severe" is often used to describe intensity of a specific event, which may be of relatively minor medical significance. "Seriousness" is the regulatory definition supplied above.*

##### *12.2 Causality*

The relationship of each adverse event to the trial medication must be determined by a medically qualified individual according to the following definitions:

**Probably related:** The adverse event follows a reasonable temporal sequence from trial vaccine administration. The event is most reasonably attributed to the study vaccination even if one or more other causes could also be reasonably attributed.

**Not probably related:** The adverse event is most reasonably attributed to the participant's underlying clinical state or by other modes of therapy administered to the participant.

##### *12.3 Safety reporting window*

Safety reporting for the trial will commence once the first participant is enrolled and will end 21 days after the last participant has received the first dose of a study vaccine for SAEs and Adverse Events of Special Interest (AESI)s.

For individual participants, the reporting period begins when they are enrolled during visit 1 and ends 21 days after the first dose of vaccine for SAEs and AESIs.

All adverse events (AEs) that result in a participants' withdrawal from the study will be followed up until a satisfactory resolution occurs, or until a non-study related causality is assigned (if the participant consents to this).

##### *12.4 Procedures for Recording Adverse Events and Vaccine Reactions*

All SAEs, medically-attended AEs, and AEs resulting in withdrawal occurring during the trial period (21 days post randomisation) will be recorded in the CRF. In addition, any solicited or unsolicited AE occurring on the day of vaccination with any COVID-19 or seasonal influenza vaccine and in the following 7 days after vaccination will also be recorded on the CRF. AEs will be recorded regardless of whether attributed to trial vaccination or not.

###### *12.4.1 Solicited adverse events following vaccination (SEFV)*

The following AEFVs will be collected by means of an electronic diary card for the 7 days after vaccination:

- Local injection site reactions:
    - Pain
    - Swelling/Induration
    - Erythema
- Systemic reaction:
    - Temperature
    - Headache
    - Fatigue
    - Chills
    - Myalgia
    - Joint pain
    - Nausea/vomiting
    - Diarrhoea

At each vaccine visit, participants will be provided with standardised instructions on how to measure temperature, redness, swelling and induration in millimetres using a tape measure, thermometer and memory aid supplied to them. Participants will be asked to record any event that is thought to represent a change in their health status. Determination of severity, seriousness and causal relationship with vaccination will be the responsibility of the study doctor at each site.

Assessment of intensity of solicited AEs will occur according to the following definitions:

| **Solicited AEFV** | **Intensity Grading** | | | | |
|---|---|---|---|---|---|
| | **None** | **Mild (Grade 1)** | **Moderate (Grade 2)** | **Severe (Grade 3)** | **Potentially life threatening (Grade 4)** |
| Pain at injection site, headache, fatigue, chills, myalgia, joint pain, nausea/vomiting, diarrhoea, | Absent | Present and causing no or minimal interference with usual everyday activities. No medical treatment | Present and causing some interference with usual everyday activities. May need medical treatment | Present and causing inability to perform usual everyday activities. Requiring medical treatment or hospitalisation | Potentially life-threatening symptoms i.e. usually an emergency, requiring urgent medical treatment |
| Temperature | 38.0 – 38.4 | 38.5 – 38.9 | 39.0 - 40 | > 40 | |
| Swelling/ Induration | < 2.5cm | 2.5 - 5cm | 5.1 - 10 cm | > 10 cm | Necrosis (requiring urgent medical care) |
| Redness (Erythema) | < 2.5cm | 2.5 - 5 cm | 5.1 - 10 cm | > 10 cm | Necrosis or exfoliative dermatitis (requiring urgent medical care) |

##### *12.4.2 Unsolicited adverse events following vaccination (AEFV)*

All local and systemic AEs occurring in the 21 days following each vaccination observed by a study investigator or reported by the participant, whether attributable to the study medication or not, will be recorded in electronic diaries or the study database. All AEs that result in a participants' withdrawal from the study will be followed up until a satisfactory resolution occurs, or until a non-study related causality is assigned (if the participant consents to this).

The intensity of unsolicited AEs will be graded as follows:

- **Mild:** Symptoms are easily tolerated and do not interfere with daily activities
- **Moderate:** Enough discomfort to cause some interference with daily activities
- **Severe:** Symptoms that prevent normal, everyday activities
- **Potentially life-threatening**: Symptoms requiring urgent medical intervention to prevent death or severe disability.

###### *12.4.3 Ascertainment of SAEs, AESI & Medical attendance*

Participants will be sent an email/text survey (and up to 2 reminders and a follow-up with a phone call if required) 21 days after COVID-19 immunisation to ascertain any event that may be considered a SAE, AESI or medical attendance. Participants will also be asked to provide the contact details of their regular doctor who will be sent a letter 21 days post-randomisation explaining the study and asking them to report any medical attendance and potential SAEs recorded within their medical record or MyHealth Record. A wallet card with the contact details for the study will also be provided to participants at enrolment instructing them to contact the study team in the event of hospital admission or the development of COVID-19.

The participant's medicare number will be recorded (as permitted by the participant) to allow for monitoring of hospital attendance to allow the study team to interrogate local hospital's electronic medical records. An investigator will obtain information from the admitting hospital to enable reporting to the Ethics Committee and if applicable, to the TGA.

The following information will be recorded for all medical attendances: description, date of onset and end date, seriousness, severity, assessment of relatedness to trial vaccination, and action taken. All AEs will be graded as mild, moderate, or severe by the investigator. Assessments of causal relationship of AEs with a study vaccine will be made for all AEs as defined in section 12.2. All reported AEs considered to be related to the study medication as judged by a medically qualified investigator or the sponsor will be followed until resolution, or until the condition is considered stable. Follow-up information should be provided as necessary.

It will be left to the investigator's clinical judgment whether or not an AE is of sufficient severity to require the participant's removal from the study. A participant may also be voluntarily withdrawn from the study due to what they perceive as an intolerable AE. If either of these occurs, the participant must undergo an end of study assessment and be given appropriate care under medical supervision until symptoms cease or the condition becomes stable.

##### *12.5 Reporting Procedures for Serious Adverse Events*

All SAEs must be reported to the Sponsor within 24 hours of study staff awareness of the SAE. An initial SAE report form must be completed with as much information as is available at the time and signed by the Principal Investigator or delegate. Or as specified in the Sponsor SOP. Additional and further requested information (follow-up until resolution or stabilisation of the event, or corrections to the original case) will be detailed on a new SAE report form and sent to the Sponsor within an appropriate time frame as specified in the Sponsor SOP.

##### *12.6 Regulatory Requirements*

The sponsor is responsible for the ongoing safety evaluation of the investigational product. The sponsor maintains overall responsibility for generating safety communications generated through feedback from the independent Data Safety Monitoring Committee. The sponsor will adhere to the reporting requirements detailed in the current version of the NHMRC safety reporting guidelines, the sponsor SOP and in line with the local regulatory guidelines.

The sponsor will provide an annual safety report to the approving HREC with a clear summary of the evolving safety profile of the trial. Additional information or individual safety reports will be provided to the HREC as specified by local guidelines.

The sponsor will notify the TGA, HREC and Investigators of all significant safety issues that adversely affect the safety of participants or materially impact on the continued ethical acceptability or conduct of the trial. The Sponsor will report all unexpected or serious adverse reactions causally related to the licensed vaccines to the appropriate authority in the relevant state.

##### *12.7 Expectedness*

Expectedness will be determined according to the Summary of Product Characteristics for each vaccine administered.

##### *12.8 SUSAR Reporting*

All SUSARs will be reported by the Sponsor to the TGA. For fatal and life-threatening SUSARS, this will be done immediately but no later than 7 calendar days after the Site Study Team is first aware of the reaction. Any additional relevant information will be reported within 8 calendar days of the initial report. All other SUSARs will be reported within 15 calendar days of being made aware of the event.

##### *12.9 Adverse events of special interest (AESI)*

| | |
|---|---|
| Immunologic | Anaphylaxis (serious allergic reaction) |

| | |
|---|---|
| Neurological | Isolated anosmia/ageusia in the absence of COVID-19<br>Guillain-Barre Syndrome<br>Acute disseminated encephalomyelitis<br>Aseptic meningitis<br>Meningoencephalitis<br>Peripheral facial nerve palsy<br>Generalised convulsion<br>Myelitis |
| Haematological | Thrombosis (excluding superficial thrombophlebitis, including line-associated)<br>Stroke<br>Thrombocytopaenia (G3 or above)<br>Eosinophilia<br>Coagulation disorder (includes coagulopathy, thrombosis, thromboembolism, internal/external bleed and stroke) |
| Cardiac | Acute cardiovascular injury (includes myocarditis, pericarditis, arrhythmias, heart failure, infarction) |
| Dermatological | Chilblain-like lesions<br>Single organ cutaneous vasculitis<br>Erythema multiforme<br>Alopecia |
| Gastrointestinal | Acute liver injury |
| Respiratory | ARDS |
| Renal | Acute kidney injury |
| Other | COVID-19 disease<br>SARS-CoV-2 positivity on a validated test |

##### *12.10 Data Safety Monitoring Committee (DSMC)*

An Data and Safety Monitoring Committee (DSMC) will be appointed to provide safety oversight. The DSMC will have an advisory role as outlined in a DSMC Charter. The DSMC will be notified of all SAEs in accordance with the DSMC charter and will undertake a timely review of these as per the Charter. In addition any Adverse Events that the CPI considers to be of safety concern to the study will be reported to the DSMC within one working day of discovery. The DSMC will inform the CPI immediately to recommend termination of the study if deemed necessary following an SAR. The DSMC will meet at least three times per year to monitor and review accumulating safety reports and will make recommendations to the CPI on whether there are any ethical or safety reasons why the trial should not continue or should be modified prior to continuation. The DSMC will consider:

- Occurrence and nature of adverse events
- Whether additional information on adverse events is required
- Consider taking appropriate action where necessary to halt the trial
- Act / advise on incidents occurring between meetings that require rapid assessment (e.g. SUSARs)

The details of the frequent interim analyses are outlined in this protocol with further detail to be included in a statistical analysis plan. Whenever an interim analysis reports inferiority, or non-inferiority with respect to the primary end-point this is termed a Statistical Trigger. At any given interim analysis, a Statistical Trigger may be reached for all participants or for one or more strata . A Statistical Trigger is reviewed by the DSMC as specified in the detailed statistical analysis plan (SAP) and the DSMC charter. When a Statistical Trigger is confirmed as having been reached by the DSMC and where no compelling reason exists not to reach a conclusion regarding that question the result that has led to a Statistical Trigger will be specified to be a stratum or trial Conclusion and potentially a Public Disclosure of the result. A Statistical Trigger can be considered as a mathematical threshold. Following any interim analyses the DSMC will make recommendations to the CPI on whether there are any ethical or safety reasons why the trial should not continue.

As noted earlier, the first interim analysis of data pertaining to the primary endpoint will be conducted by the trial statistician when 200 participants enrolled to a single COVID-19 brand have completed the first 7-day follow-up period. Following the interim analysis the DSMC will make recommendations to the CPI on whether there are any ethical or safety reasons why the trial should not continue. Further interim analyses will be conducted by the trial statistician as outlined in Section 13.

The DSMC may request to be unblinded to the treatment allocation of individual participants and/or to have additional blinded or unblinded analyses (beyond those specified in the SAP) performed by the trial statistician.

The DSMC will have a minimum of two independent advisors and a statistician. The Chair of the DSMC will be contacted for advice and independent review in the following situations:

- Following any SUSAR
- A Statistical Trigger met (see Section 13)
- Any other situation where the Investigator feels independent advice or review is important

#### 13. Statistical Methods

Inference for the trial will be derived from the accruing data at recurrent interim analyses. Generalised linear models will incorporate relevant aspects of the design, accounting for variation by site and time. The model for the primary analysis will be logistic regression (or equivalent) with weakly informative priors to compute the joint posterior distributions that characterising the change in the propensity of solicited adverse events of at least moderate severity, following COVID-19 vaccine co-administered with seasonal influenza vaccine relative to COVID-19 vaccine co-administered with placebo, for each SIV vaccine, for each COVID-19 vaccine brand, and for each COVID-19 vaccine dose if differential effects are apparent. The priors used are such that for sample sizes in excess of 50 per arm, these priors produce results that are practically identical to frequentist maximum likelihood estimates.

The posterior distributions will be used to evaluate decision points for declaring inferiority and non-inferiority. There is no broadly accepted minimum clinically important difference for the frequency of adverse reaction following vaccination. An absolute 15% non-inferiority margin is used for the trial, and corresponds to the expected rate of moderate or severe reaction when SIV is administered alone, and about one additional moderate or severe reaction for every 7 vaccinated people. However it will be for policy-makers to determine whether any observed increase is acceptable. The reference thresholds used to control when decisions are made will be refined based on further trial simulations under a range of scenarios.

A comprehensive statistical analysis plan (SAP) will be prepared prior to the first interim analysis. Development of the SAP will only be done by personnel who are blinded to the participant data.

#### 14. Data Management

Data will be collected into a secure local instance of the Sydney Local Health District (SLHD) REDCap database. Data will be entered into an electronic Case Report Form (eCRF) developed in accordance with the protocol. Data entry will be performed by trained study staff, apart from electronic patient reported outcome data which will be directly entered by the participant. Information will be recorded in the eCRF to accurately reflect the source data. All users will have appropriate permissions defined by their user role type which is delegated by the Sponsor and Site Investigator.

Data entry and data management will be coordinated by the SLHD site. The study coordinator, or delegate, will perform regular and timely validation of data, queries and corrections. Any common patterns of errors will be reported back to participating sites. Missing data will be minimized through a clear and comprehensive data dictionary with online data entry including logic checks. Procedures to ensure data quality and protocol will also include: a dictionary to define the data to be collected on the CRF/eCRF; start-up meetings to ensure consistency in procedures; site induction before activation of the site to enrolment, with specific training for clinical staff; and development and dissemination of guidance materials.

##### *14.1 Record Keeping*

Individual participant data and records will be held securely and in the strictest confidence by the participating site and by the study coordinating centre, as required and permitted by law. On all study-specific documents, other than the signed consent form, the participant will be referred to by a unique study-specific number and/or code in any central database, not by name or other identifying information. Information linking the participant's medical data to database material will be maintained securely and accessible only by staff at the relevant site. The key to re-identify participants from their study identifier will only be accessible to staff at the local site.

##### *14.2 Data Retention and Disposal*

In the case of closure of the study, the sponsor will retain an identical replica of the platform database for 15 years or longer, as is required by the approving regulatory authorities.

Any paper documents will be stored at participating sites in locked cupboards whilst the platform is live. These will be archived until 15 years after the end of the study/publication, whichever is later.

Any electronic data recorded on the platform database will also be archived according to the standard procedures of the Sponsor relating to archiving of electronic data. Documents will be destroyed as per the sponsor's Information Retention & Disposal Policy and with authorisation from the Coordinating Principal Investigator.

##### *14.3 Data Security*

Study participant data will be identified by a unique study ID number on all study documentation, including the electronic database.  All documents will be stored securely and will only be accessible by project staff and authorised personnel. This study will comply with all laws pertaining to data protection. Data will be anonymised as soon as it is practical to do so.

In order to administer electronic patient reported outcomes while ensuring participant's anonymity, access to participant contact details will be limited to minimum authorised research staff. Minimum contact data will be collected to ensure follow up can be completed, including name, email address and mobile phone number.

##### *14.4 Data Collected*

| Variable | Timepoint | Justification |
|---|---|---|

| | | |
|---|---|---|
| Demographic information, including date of birth, sex, and ethnicity/ Indigenous status, occupation | Enrolment | To permit an adequate description of the study population.<br><br>Factors like age and sex plausibly influence immune responses and reactogenicity to vaccination. Age is a pre-specified stratifying variable for immunogenicity analyses. Self-reported ethnicity, including Indigenous status, may also influence vaccine responses, both as a consequence of genetic factors, and also as a consequence of socio environmental factors and comorbidities. Demographic details can also help confirm accurate linkage of healthcare and administrative data |
| Relevant clinical history relating to potential predictors of COVID-19 disease or modifiers of vaccine effects, including any concomitant medications | Enrolment | To permit an adequate description of the study population.<br><br>The presence of underlying comorbidities plausibly influence immune responses to vaccination, in particular those which are clearly immunocompromising like primary and acquired immunodeficiencies, but also other acute and chronic conditions like renal and liver impairment. Likewise, medications may directly affect immune responses (like steroids and other immune modulating treatments), or may affect reactogenicity of vaccines. |
| Anthropometric measures including weight/ BMI | Enrolment | Overweight and obesity are plausibly associated with reduced vaccine responses, both as a consequence of the direct effects on the immune system, and because obesity is a risk factor for inadvertent subcutaneous administration of intramuscular vaccines |
| Vaccine received, date, batch number, body site of administration, arm of | Each vaccine administered | Capture of the vaccine administered and date of receipt will allow assessments of comparative immunogenicity, effectiveness and safety across vaccine types. The date of administration will enable assessments of the durability vaccine effects and the temporal relationship of any |

| | | |
|---|---|---|
| COVID-19 vaccine and SIV | | adverse events. The batch number, if available, will enable us to evaluate for any batch-specific vaccine effects. Record of the site of administration may be informative for attribution of local reactions to vaccine, and because a significant part of the vaccine immune response occurs in the draining lymph nodes, site of administration may be relevant for understanding any interaction/interference from co-administered vaccines, and booster responses to subsequent vaccination. |
| Participant identifiers (phone number, address and Medicare number) to be accessible by the site only | Enrolment | Each site will capture and retain identifying information to enable proof-of-identity (in case of future safety issues and to ensure trial integrity), to enable contact by the study team for study-related procedures or invitation to participate in future sub-studies, and to enable individual linkage to relevant data contained in other healthcare and administrative datasets. Identifying information will not be made accessible to those outside of the site. |
| Self-reported adverse reactions: pain at injection site, headache, fatigue, chills, myalgia, joint pain, nausea/vomiting, diarrhoea, swelling/ induration, redness (Erythema) | Daily for 7 days after each vaccination | Primary endpoint. To determine if the incidence of moderate (or worse) solicited adverse reactions occurring up to 7 days after co-administration of COVID-19 and influenza vaccines is non-inferior to co-administration COVID-19 and placebo. |
| Medical attendance or hospitalisation post immunisation and change in medication (including any additional vaccines) | Up to 21 days post randomisation | While the mild and even moderate reactions to vaccination are to be expected and are unlikely to materially affect their risk-benefit profile, reactions that result in medical attendance or hospitalisation are usually severe and possibly serious, and affect the risk-benefit profile. Severe events may plausibly occur at any time |

| | | |
|---|---|---|
| | | after vaccination and differentiating events that are attributable to vaccination from those that are merely coincidental is challenging, but those occurring close to vaccination (up to 7 days afterwards) are more likely to warrant particular attention than those that are more distant. Requesting self-reports on day 21 is most likely to ensure the accuracy of the collected data.<br><br>Changes in medication received including additional vaccines may affect the participants response to the study vaccines in terms of reactogenicity and immunogenicity. |
| Self-reported COVID-19 testing, approximate date and location of any COVID-19 laboratory test and laboratory-confirmed diagnosis | Every month for up to 6 months | While COVID-19 transmission in Australia is low at the time of commencement of the trial, it cannot be assumed that this will be the case indefinitely. The relative incidence of laboratory-proven infection is the most reliable measure of the efficacy of vaccines, and so any proven ‘breakthrough’ infection among vaccinated people is highly relevant to the overall assessment of their performance, and will supplement assessments of immunogenicity. Self-report of infection will assist research staff in confirming infection with the relevant laboratory and/or public health authority, and permit ascertainment of any genomic or sequencing data from detected strains.<br><br>Ascertaining all tests (regardless of result) will help ensure more complete ascertainment, and will also permit a description of the microbiological follow-up of study participants and an assessment of differential follow-up among different subgroups. |
| Self-reported presence and severity of any symptoms related to COVID-19 infection, including | Every month for up to 6 months | Among those with laboratory-confirmed COVID-19 infection, understanding the severity of infection is relevant to an assessment of the effectiveness of the vaccine. Many vaccines are more highly protective against severe rather |

| | | |
|---|---|---|
| management received for COVID-19 infection, including need for medical attendance or hospitalisation | | than mild or asymptomatic infection. In general, for this purpose, severity can be pragmatically assessed by whether there were any respiratory symptoms or not, whether and symptoms resulted in a medical attendance, whether they resulted in hospitalisation and/or oxygen therapy, whether they resulted in intensive care admission, and whether they resulted in death. |
| Self-reported presence of influenza-like-illness (ILI) or laboratory confirmed influenza | Every month for up to 6 months | Ascertainment of ILI and laboratory-confirmed influenza infections will enable us to assess the comparative effectiveness (and possible inferiority) of co-administering SIV with COVID-19 versus administration 7-14 days after COVID-19. Measurement of haemagglutinating antibodies, by itself, is an imperfect predictor of clinical effectiveness. |

#### 15. Quality Assurance Procedures

This study will be conducted in accordance with the current approved protocol, ICH GCP, the National Statement on Ethical Conduct in Human Research, and relevant regulations. A risk assessment will identify and define the data and processes critical to participant safety and data quality, identifying the risks, and creating processes to minimize them. Indicators and thresholds will be set that will trigger an investigation and/or corrective action. This risk-based approach will be used by the Project Manager to develop a monitoring plan for approval by the Steering Committee; it will detail any obligations expected of sites to assist the Sponsor in monitoring including hosting site visits, providing information for remote monitoring, or putting procedures in place to monitor the study internally. Other procedures to ensure data quality include:

- A detailed data dictionary will define the data to be collected on the case report form;
- Quality checks will be built into the data management system and there will be quality checks of critical data points entered into the CRFs to ensure standardization and validity of the data collected;
- Data queries may be generated, depending on resource availability. Any information that is not available for the investigator will not be considered as missing. No assumptions will be made for missing data.

### 16. Protocol Deviations/Violations

#### *16.1 Protocol deviation*

A protocol deviation is any change, divergence, or departure from the study design or procedures of a research protocol that does not increase risk or decrease benefit or; does not have a significant effect on the subject's safety or welfare; and/or on the integrity of the data. Prospective, planned deviations or waivers to the protocol are not permitted e.g. it is not acceptable to enrol a subject if they do not meet the eligibility criteria or restrictions specified in the trial protocol.

Accidental protocol deviations can happen at any time. Protocol deviations will be documented in the CRF and a log of protocol deviations will be maintained by site. This log will be reviewed on a regular basis by the Sponsor to assist with monitoring protocol compliance. Deviations from the protocol which are found to frequently recur are not acceptable, will require immediate action and could potentially be classified as a serious breach.

#### *16.2 Serious Breaches*

##### *16.2.1 Suspected Breach*

A suspected breach is defined as:
A report that is judged by the reporter as a possible serious breach but has yet to be formally confirmed as a serious breach by the sponsor.

Suspected breaches will be treated as a serious breach until deemed otherwise by the trial Sponsor.

##### *16.2.2 Serious Breach*

A serious breach is defined as:
A breach of Good Clinical Practice or the protocol that is likely to affect to a significant degree: a) The safety or rights of a trial participant, or b) The reliability and robustness of the data generated in the clinical trial. Note: this guidance's definition of serious breach differs from the definition in the Australian Code for the Responsible Conduct of Research and is about deviations from the requirements of Good Clinical Practice or the clinical trials protocol.

In the event that a serious breach is suspected, the Sponsor must be contacted within 1 working day. In collaboration with the CPI, the serious breach will be reviewed and reported (if required) by the Sponsor in accordance with the current NHMRC guidance *Reporting of Serious Breaches of Good Clinical Practice (GCP) or the Protocol for Trials Involving Therapeutic Goods.*

### 17. Ethical and Regulatory Considerations

#### *17.1 Declaration of Helsinki and Guidelines for Good Clinical Practice*

Study investigators and all study staff will ensure that this trial is conducted in accordance with the principles of the Declaration of Helsinki and the *International Council for Harmonisation (ICH) Guidelines for Good Clinical Practice* (CPMP/ICH/135/95) July 1996.

#### *17.2 Approvals*

The protocol, participant information sheet and informed consent forms, and any proposed advertising material will be submitted to one or more Human Research Ethics Committees (HREC), and as required, to any regulatory authorities. Written approval to commence the study will be required from all relevant ethical and regulatory bodies at participating sites. No changes to any sections of the study protocol, or participant information sheet and informed consent form (unless trivial, for example a change in contact phone number) will be made without prior written approval of the approving HREC. Any trivial changes will be reported and ratified by the HREC at the earliest opportunity.

#### *17.3 Potential Risk for the Participant*

##### *17.3.1 Confidentiality*

Participation in this study involves the potential risks of accidental breach of confidentiality of the recorded information and breach of the privacy of the participants. This will be minimised by: removing direct participant identifiers (i.e. names, Medicare numbers, medical record numbers) from the information stored for the study and used for analysis, securing and limiting access to linking codes assigned to the study information, limiting access to information contained within the study to authorised research staff.

##### *17.3.2 Coadministration of COVID-19 and Influenza vaccine*

There are no specific indicators to suggest co-administration of TGA approved seasonal influenza and COVID-19 vaccines is likely to be ineffective or harmful. However, there are no known studies demonstrating the reactogenicity and immunogenicity of simultaneous administration of Comirnaty or COVID-19 vaccine AstraZeneca with influenza vaccines. Systemic adverse reactions and injection site reactions occur in approximately 40 - 60% of COVID-19 vaccine recipients and less than 10% of SIV vaccine recipients in the few days following vaccination when given alone.

The Australian Immunisation Handbook advises that precautions should be taken for individuals with egg allergy. Most people with egg allergy can be safely vaccinated with egg-based influenza vaccines due to low concentrations of ovalbumin, however longer waiting may be recommended if anxiety is apparent.

It is plausible that participants will experience increased adverse reactions and/or a poorer immune response with coadministration of seasonal influenza vaccines with COVID-19 vaccines. This will be explained to participants within the PISIC process. Adverse reactions will be monitored daily for 7 days after each vaccination.

On May 27th 2021, the Chief Medical Officer of Australia announced that the 14 day window previously recommended between administration of COVID-19 vaccines and other vaccines could be waived for residents and staff in residential aged care. With this recommendation came assurance that the effectiveness of co-administration of influenza and COVID-19 vaccines will not be impacted.

On June 9th, ATAGI changed the 14 day window to 7 days.

###### *17.3.3 Phlebotomy*

For participants who opt-in to providing blood samples for the study, blood sampling may cause temporary discomfort but the amount of blood to be taken is unlikely to cause symptoms. The maximum volume of blood to be collected at a single visit is 20mL and is unlikely to cause or precipitate anaemia. There may be slight bruising at the site where the blood was taken, however this is unlikely to persist beyond a few days. Venepuncture may induce fainting or a feeling of ‘light-headedness’ due to a vasovagal response.

##### *17.4 Potential Benefit for the Participant*

Participants in the study receiving a complete schedule of COVID-19 vaccine and influenza vaccine should have a lower risk of COVID-19 disease and influenza disease than unimmunised individuals.

It is hoped that the information gained from this study will contribute to a reduction in deaths and suffering from COVID-19 disease and influenza disease through the generation and timely dissemination of high-quality evidence to inform immunisation strategies.

##### *17.5 Declarations of Interest*

Study investigators declare have no interests to declare.

##### *17.6 Funding*

This study will be funded by Sydney Local Health District and Snow Medical Research Foundation

##### *17.7 Insurance*

Trial indemnity is provided by the study sponsor.

##### *17.8 Partnering with consumers*

The study was designed in consultation with the National COVID-19 Community Reference Group (CRG) who had input into the design and reviewed the participant information sheet and consent form, as well as the electronic diary card. On-going involvement will be stewarded by community representative Mr Mitch Messer, including where necessary via additional meetings with the CRG.

#### 18. Reporting and Publication Policy

##### *18.1 Reporting*

Each participating site will comply with local reporting requirements, as specified by that site's institution. Should the study be terminated, all relevant local ethical and regulatory bodies will be informed within 90 days after the end of data collection.

##### *18.2 Communication of Results*

Study results will be communicated by presentation and publication.

##### *18.3 Publication Policy*

The study investigators will, as far as possible, make the protocol, statistical analysis plans, and non-identifying participant-level data available in order to allow independent scientific scrutiny and validation of any published results. All Investigators will have the opportunity to review publications (e.g., manuscripts, abstracts, oral/slide presentations, book chapters) prior to submission.

Authorship will be determined in line with the Uniform Requirements for Manuscripts Submitted to Biomedical Journals published by the International Committee of Medical Journal Editors. In summary, authorship will be limited to those who have:

- Contributed substantially to the conception and design of the study; or the acquisition, analysis or interpretation of data for the work; AND
- Drafted the work or revised it critically for important intellectual content; AND
- Provided final approval of the version to be published; AND agreed to be accountable for all aspects of the work in ensuring that questions related to the accuracy or integrity of any part of the work are appropriately investigated and resolved.

Acquisition of funding or general supervision of the research group alone will not constitute grounds for authorship. The final decision on authorship of any publication will be the responsibility of the CPI. If the study investigators agree to provide any pre-publication versions of presentations or manuscripts to a commercial organisation, the commercial organisation will have no authority to prevent, inhibit, or modify presentation or publication.

21. Australian Technical Advisory Group on Immunisation (ATAGI. Australian Immunisation Handbook [Internet]. Australian Government Department of Health, Canberra; 2018. Available from: immunisationhandbook.health.gov.au.

22. Public Health England. Immunisation against infectious disease (Green Book) [Internet]. Department of Health (UK), London; Available from: https://www.gov.uk/government/collections/immunisation-against-infectious-disease-the-green-book#the-green-book

#### 16. Amendment History

| **Amendment no.** | **Protocol version no.** | **Date issued** | **Details of changes made** |
|---|---|---|---|
| N/A | N/A | N/A | N/A |

| | | | |
|---|---|---|---|
| 1 | 2 | 1/11/2021 | Addition of boosters, changes to ATAGI recommendation (October 2021) |
| 2 | 3 | | Update background section with ATAGI recommendation describing additional SIV.<br>Additional exclusion criteria of “Receipt of SIV within the preceding 6 months” |
| 3 | 4 | | Addition of two additional influenza vaccines:<br>1. Sanofi Vaxigrip Tetra<br>2. Sanofi Fluzone High-Dose Quadrivalent<br>Modification of the definition of SAE to specify inclusion of ‘unplanned hospitalisation’ |
