## Appendix B: Daily diary card for "A single blinded, phase IV, adaptive randomised control trial to evaluate the safety of coadministration of seasonal influenza and COVID-19 vaccines (The FluVID study)"

Please complete the survey below.

Thank you!

Date ______________________

### LEFT ARM

Pain at the injection site

- ◯ No pain
- ◯ Mild pain (with no impairment to usual activities)
- ◯ Moderate pain (with some impairment to usual activities)
- ◯ Severe pain (unable to perform usual activities)
- ◯ Life-threatening

Measured swelling or induration (hardening of the skin) at the injection site

- ◯ < 2.5cm
- ◯ 2.5 - 5cm
- ◯ 5.1 - 10cm
- ◯ > 10cm
- ◯ Required urgent medical care

(Please use the tape measure provided to measure any area of swelling and/or hardness at the injection site of the vaccine in cm.)

Measured area of redness at the injection site

- ◯ < 2.5cm
- ◯ 2.5 - 5cm
- ◯ 5.1 - 10cm
- ◯ > 10cm
- ◯ Required urgent medical care

(Please use the tape measure provided to measure any area of redness (erythema) at the injection site of the vaccine in cm)

### RIGHT ARM

Pain at the injection site

- ◯ No pain
- ◯ Mild pain (with no impairment to usual activities)
- ◯ Moderate pain (with some impairment to usual activities)
- ◯ Severe pain (unable to perform usual activities)
- ◯ Life-threatening (required urgent medical treatment)

Measured swelling or induration (hardening of the skin) at the injection site

- ◯ < 2.5cm
- ◯ 2.5 - 5cm
- ◯ 5.1 - 10cm
- ◯ > 10cm
- ◯ Required urgent medical care

(Please use the tape measure provided to measure any area of swelling and/or hardness at the injection site of the vaccine in cm)



REDCap

| | |
|---|---|
| Measured area of redness at the injection site | ◯ < 2.5cm<br>◯ 2.5 - 5cm<br>◯ 5.1 - 10cm<br>◯ > 10cm<br>◯ Required urgent medical care<br>(Please use the tape measure provided to measure any area of redness (erythema) at the injection site of the vaccine in cm) |

### GENERAL SYMPTOMS YOU EXPERIENCED IN THE LAST 24 HOURS

| | |
|---|---|
| Have you had a fever? | ◯ Yes<br>◯ No |
| What was the highest temperature you recorded? | ◯ 38.0 - 38.4<br>◯ 38.5 - 38.9<br>◯ 39.0 - 40<br>◯ > 40<br>(in degrees Celsius) |
| Headaches | ◯ No headache<br>◯ Mild headache (with no impairment to usual activities)<br>◯ Moderate headache (with some impairment to usual activities)<br>◯ Severe headache (unable to perform usual activities)<br>◯ Life-threatening (required urgent medical treatment) |
| Fatigue (tiredness) | ◯ No fatigue<br>◯ Mild fatigue (with no impairment to usual activities)<br>◯ Moderate fatigue (with some impairment to usual activities)<br>◯ Severe fatigue (unable to perform usual activities)<br>◯ Life-threatening (required urgent medical treatment) |
| Chills (shivering) | ◯ No chills<br>◯ Mild chills (with no impairment to usual activities)<br>◯ Moderate chills(with some impairment to usual activities)<br>◯ Severe chills (unable to perform usual activities)<br>◯ Life-threatening (required urgent medical treatment) |
| Widespread muscle pain | ◯ No muscle pain<br>◯ Mild muscle pain (with no impairment to usual activities)<br>◯ Moderate muscle pain (with some impairment to usual activities)<br>◯ Severe muscle pain (unable to perform usual activities)<br>◯ Life-threatening (required urgent medical treatment) |

| | |
|---|---|
| Joint pain (not including injection site) | ◯ No joint pain<br>◯ Mild joint pain (with no impairment to usual activities)<br>◯ Moderate joint pain (with some impairment to usual activities)<br>◯ Severe joint pain (unable to perform usual activities)<br>◯ Life-threatening (required urgent medical treatment) |
| Nausea/vomiting | ◯ No nausea/vomiting<br>◯ Mild nausea/vomiting(with no impairment to usual activities)<br>◯ Moderate nausea/vomiting (with some impairment to usual activities)<br>◯ Severe nausea/vomiting (unable to perform usual activities)<br>◯ Life-threatening (required urgent medical treatment) |
| Diarrhoea | ◯ No diarrhoea<br>◯ Mild diarrhoea (with no impairment to usual activities)<br>◯ Moderate diarrhoea (with some impairment to usual activities)<br>◯ Severe diarrhoea (unable to perform usual activities)<br>◯ Life-threatening (required urgent medical treatment) |

### MEDICATIONS AND MEDICAL CARE IN THE LAST 24 HOURS

| | |
|---|---|
| Did you take any medication for pain relief? | ◯ Yes<br>◯ No<br>(e.g. paracetamol, ibuprofen) |
| Did you take any antihistamine? | ◯ Yes<br>◯ No<br>(e.g. loratadine, cetirizine) |
| Did you take any anti-nausea medication? | ◯ Yes<br>◯ No<br>(e.g. kwells, ondanserton) |
| Did you seek any medical advice? | ◯ Yes<br>◯ No |
| From who/where did you seek medical advice? | ◯ Phone advice (e.g. HealthDirect)<br>◯ Care from a GP or Aboriginal Healthcare worker (in person, telehealth, email, urgent care clinic, home visit)<br>◯ Visit to a hospital emergency department |
| In the past 24 hours, did any of the symptoms you reported cause you to miss work, study or normal daily activities? | ◯ Yes<br>◯ No |
