## Appendix C: Day 21 survey for "A single blinded, phase IV, adaptive randomised control trial to evaluate the safety of coadministration of seasonal influenza and COVID-19 vaccines (The FluVID study)"

Please complete the survey below.

Thank you!

### IN THE LAST 21-DAYS (SINCE YOU STARTED THE FLU-VID STUDY)

Did you seek medical care?

◯ Yes
◯ No

From who/where did you seek medical care?

◯ Your Doctor (GP)
◯ Hospital emergency department
◯ Admitted to hospital

Which clinic or hospital did you attend?

__________________________________

Since enrolling into the study, have you been diagnosed by a doctor with any of the following?

◯ COVID-19 infection
◯ Anaphylaxis or other serious allergic reaction
◯ Sudden loss of taste or smell
◯ Guillain-Barre syndrome or other severe inflammation of the brain, spinal cord or nervous system
◯ Bell's palsy (paralysis affecting the face)
◯ Seizure or convulsion
◯ Thrombosis or blood clot
◯ Stroke
◯ Disorder of any of the blood cells (too many or too few)
◯ Myocardial infarction (heart attack), unstable angina or acute coronary syndrome
◯ Chilblains
◯ Vasculitis or inflammation affecting the blood vessels
◯ Erythema multiforme skin rash
◯ Alopecia (sudden unexplained hair loss)
◯ Sudden severe inflammation of the lung, liver or kidney

If other, please specify

__________________________________________

Did you take time off work, study or usual activities because of any of the symptoms you have reported for this study?

◯ Yes
◯ No

How many days?

__________________________________
