## Appendix D: Statistical analysis plan for "A single blinded, phase IV, adaptive randomised control trial to evaluate the safety of coadministration of seasonal influenza and COVID-19 vaccines (The FluVID study)"

**A single blinded, phase IV, adaptive randomised control trial to evaluate the safety of co-administration of seasonal influenza and COVID-19 vaccines - Statistical Analysis Plan**

Mark Jones

Date: 2021-10-18
Sponsor: Sydney Local Health District
HREC Number: 2021/ETH01232
Protocol No: X21-0209
ANZCTR number: ACTRN12621001063808p

#### *Revision History*

| Revision | Date | Author(s) | Description |
|---|---|---|---|
| 0.1 | 2021/08/19 | MAJ | Created SAP. |
| 0.2 | 2021/09/13 | MAJ | Updating after review 1. |
| 0.3 | 2021/10/05 | MAJ | Updating based on comments from TS. |
| 0.4 | 2021/10/18 | MAJ | Revised interim start. Generalising COVID-19 Vaccines. |

**References** **20**

#### *List of Figures*

1 CONSORT template . . . 17

### 1 Introduction

FluVID is a pragmatic Bayesian adaptive, placebo controlled randomised trial evaluating the safety of co-administering seasonal influenza vaccines (SIV) with COVID-19 vaccines in Australia.

This document broadly follows the outline per Gamble et al. (2017) and presents the general framework for the statistical design and analysis plan for the trial.

#### 1.1 Background and Rationale

The full rationale for undertaking the trial along with the trial background are explained in the protocol. In brief, the motivation for the FluVID trial stems from the common practice of simultaneously administering different vaccines for both children and adults. Co-administration of COVID-19 vaccines with seasonal influenza vaccines, if proven to be safe and effective, offers significant advantages for health systems planning. Potential benefits include reduced cost, administrative practicality, effective use of resources and increased uptake of both vaccines. Therefore, FluVID has been designed to assess reactogenicity of coadministering COVID-19 vaccines with SIV.

#### 1.2 Objectives

##### 1.2.1 Primary Objectives

To determine if the proportion of participants experiencing moderate or severe (grade 2 or above) adverse reactions up to 7 days after co-administration of SIV with COVID-19 vaccines is no more than an absolute increase of 15% above the proportion of participants that experience mod/sev AEs in participants that are coadministered saline placebo with COVID-19 vaccine.

##### 1.2.2 Secondary Objectives

1. To compare co-administration of COVID-19 vaccine with SIV versus co-administration of COVID-19 vaccine with saline placebo in terms of the occurrence of:

   - solicited adverse reactions
   - serious adverse events
   - adverse events of special interest

   7-14 days following vaccination.

2. To determine whether the effectiveness of each of the SIV and COVID-19 vaccines is no worse (risk of disease increases by no more than 15% in relative terms) when they are co-administered than when SIV is administered 7-14 days after COVID-19 vaccine.

For the second secondary objective, we will examine the effectiveness of SIV and COVID-19 when delivered together relative to delivering them independently (or with placebo). That is, the goal is to measure SIV efficacy via the occurrence of influenza like illness or laboratory confirmed influenza up to 6 months after visit 1 (for the cohort that received COVID-19+SIV) and compare this with the analogous measure for the cohort that received SIV (alone) at visit 2. For the COVID-19 vaccine efficacy we compare the response in the cohort that received COVID-19 and SIV combined with the analogous measure from the cohort that receives COVID+19 combined with placebo at visit 1.

Further detail of the specific outcomes of interest are listed in the protocol and under the Outcomes section of this document.

### 2 *Structure of Trial*

#### 2.1 Design

The study design is a Bayesian sequential multi-arm placebo-controlled trial with adaptive sample size, incorporating stopping rules for both inferiority and non-inferiority for each COVID-19 vaccination brand/dose pair.

The trial aims to compare co-administration of SIV with COVID-19 (at any dose) versus co-administration of placebo with COVID-19 vaccine (at the same dose). The trial requires that participants make two visits to the clinic over the course of their involvement. At the first visit, they recieve a co-administered dose and at the second they receive either a SIV or placebo. This ensures that all participants receive the SIV vaccine. Initially, Pfizer and Astra Zeneca branded COVID-19 vaccines were available. However, the situation is fluid and as new COVID-19 vaccines come online they will be incorporated into the trial. Irrespective of the vaccines brands used, throughout this document we refer to co-administration of SIV with any COVID-19 vaccine as the *investigational arm* and co-administration of placebo with the same COVID-19 vaccine brand (at either dose) as the *control arm*. The primary outcome is the occurrence of any moderate or severe adverse events within a 7-day follow up period.

The design includes sequential analyses that will start when any single COVID-19 brand has 200 participants that have reached their 7-day follow up after the first visit. Subsequent analyses will occur at increments of 100 new participants reaching their 7-day follow up. At each analysis, pre-specified stopping rules are evaluated and reported to the DSMC for review. Based on current resources, the maximum sample size has been set at 1000 participants. The trial can stopped before reaching this sample size if decisions are reached on all arms. Additionally, the trial can be extended if additional resources become available and the need arises.

#### 2.2 Outcomes

Table 2 shows the study outcomes.

Table 2: Study outcomes and representation

| Outcome | Type | ID | Description | Representation |
|---|---|---|---|---|
| Primary | Safety | PS1 | Any solicited adverse reaction of severity grade 2-4 occurring up to 7 days following administration of influenza vaccine or placebo administered with COVID-19 vaccine (visit 1) | Binary |
| Secondary | Safety | SS1 | Solicited *local* reactions, up to 7 days after visit 1 and visit 2 | Ordinal |
| | Safety | SS2 | Solicited *systemic* reactions, up to 7 days after visit 1 and visit 2 | Ordinal |
| | Safety | SS3 | Any serious adverse event up to 21 days after COVID-19 immunisation (visit 1) | Binary |
| | Safety | SS4 | Any adverse events of special interest up to 21 days after COVID-19 immunisation (visit 1) | Binary |
| | Safety | SS5 | Any medical attendance up to 21 days after COVID-19 immunisation (visit 1) | Binary |
| | Safety | SS6 | Any days off work for *employed* participants up to 7 days, and up to 21 days after visit 1 and visit 2 | Binary |
| | Effectiveness | SE1 | Laboratory-confirmed COVID-19 infection up to 6 months after completion of primary immunisation (visit 1) | Binary |
| | Effectiveness | SE2 | Laboratory-confirmed influenza infection up to 6 months after completion of primary immunisation (visit 1) | Binary |

##### 2.3 Randomisation

Participants are both the experimental and observational units. Treatments (investigational or control arms) are randomised (1:1) stratified by:

- site (initially one site)
- brand (initially two brands)
- dose (initially two doses)
- SIV type

The randomisation procedure uses permuted blocks with sizes of 4, 6 and 8 (with equal probability) and was implemented by MJ. Research staff assign participants via a Redcap database at the time of participant enrollment. The random number seeds, code and master list are held on restricted access network folders currently only accessible to MJ. Stratification will be accounted for in the analyses either by random or fixed effects as necessary, see Fleiss (1986) and Piantadosi (2017).

##### 2.4 Sample size

Under a Bayesian approach, pre-specification of the sample size is not as important as it is in a frequentist study (Berry et al., 2010; Kruschke and Liddell, 2018), but there is still much debate on the topic, e.g. Spiegelhalter et al. (2004) page 199. The sample size for FluVID is adaptive and is governed by pre-specified rules. This means that the sample size is a random variable and is unknown prior to being observed. We have set a maximum sample size of 1000 based on the availability of resources, practicality and initial trial simulation. However, as the sample size is adaptive, the trial may stop before reaching 1000 enrolments. For example, current simulations across a range of scenarios suggest an expected sample size around 700, i.e. if two COVID-19 brands were in use. Finally, if additional resources become available and there remain unanswered questions, we may extend trial recruitment beyond 1000.

##### 2.5 Blinding

FluVID is a placebo controlled trial with blinding of both the participants and the research staff administering the vaccinations. Both the trial statistician and DSMC will be unblinded to treatment allocation and cannot be involved in variations to the trial design subsequent to unblinding. No personel involved in the development of this statistical analysis plan has reviewed any of the data.

When publicly reporting the results of a statistical decision for a COVID-19 vaccination brand/dose pair, only information relevant to the brand/dose pair on which the decision has been made will be disclosed.

### 3 *Statistical Modelling*

#### 3.1 Analysis Population

The primary analysis population will include all participants that were randomised and reached 7 days after randomisation with their primary endpoint status either known or known to be missing. The day that the participant is randomised (regardless of the time of day) is treated as day 1 and the data collected up to midnight on the end of day 7 constitutes the 7 days of follow up data. This population will be used for all outcomes and will follow the intention-to-treat (ITT) principle. Specifically, all randomised patients will be included and analysed according to the treatment arm to which they were initially assigned irrespective of deviations from the treatment or any other protocol deviations. Data for participants who have withdrawn will be included in the primary analysis population up to the time they withdrew. Participants that have reached the 7-day follow up, but for whom information has not yet been collected will be treated as missing until the data has been entered. Participants who have been randomised, but have not yet reached the 7 day follow up, will be excluded from analyses.

A secondary analysis population will include all participants who are randomised to at least one of the interventions. This analysis set will follow the per-protocol (PP) definition with randomised patients included in the analysis only if they received their allocated treatment and no protocol deviations occurred prior to the primary endpoint.

Additional analysis populations may be of interest to inform secondary and sensitivity analyses. For example, outcome `SS6` (Table 2) is only applicable for participants that are employed at baseline. These will be specified and reported as needed.

#### 3.2 Interim analyses

Interim analyses have been specified to start when 200 participants have accrued to a single COVID-19 brand (and reached their 7 day follow up) and occur for every 100 enrollments thereafter. With permission from the DSMC, this rule may be varied depending accrual rates and availability of resources.

The interim analyses will be based on both the ITT and PP populations as defined in the Analysis Population section and decisions made only if both data sets result in consistent conclusions.

At each interim analysis, Trial Decision Criteria are evaluated and, if met, may be publicly reported. The reported results will be based on all the available data to maximally incorporate information providing more robust estimation of all included covariates. However, only the information relevant to the decision criteria that has been met will be disclosed. We will also complete an analysis that is based on a subset of the data restricted to the brand/dose pairs on which the decision was made. This latter approach is more in line with a conventional trial.

While imputation may be used to complete missing covariate values, no imputation will be undertaken on the outcome measures.

#### 3.3 Final analyses

In the event that the trial runs to 1000 participants without a decision being made, all participants will be followed up and the final analyses completed using the data sets as defined in the Analysis Population section.

For early trial decisions (including stopping the trial entirely) the exact procedure is at the discretion of the DSMC. However, indicatively, all enrolled participants belonging to the relevant arms would be followed up for the required duration and a final analysis completed using the data relating to those intervention and control arms. For example, as the trial proceeds it may become apparent that coadministration of Pfizer dose one with SIV is non-inferior to coadministration of Pfizer dose one with placebo. If the pre-specified decision threshold is met, the DSMC will likely recommend that further enrolment for Pfizer dose one is stopped, and we will follow up all participants in the Pfizer (dose one) groups (both the investigational and control arms) and complete a final analysis on this dataset. As such, under early stopping, the final analysis for this example will only use data relating to the specific brand/dose group(s) for which the decision threshold was met. The above approach may be varied at the discretion of the DSMC if concerns that public announcements of trial decisions could compromise the integrity of the trial.

##### 3.4 Baseline data

Descriptive statistics on a range of patient characteristics as defined by clinical relevance will be included with all analyses. Summaries of these variables (counts and proportions for discrete, median and inter-quartile range for continuous) will be tabulated and presented in aggregate and by intervention. No statistical tests will be run to compare groups.

##### 3.5 Analysis of primary outcome

The primary outcome will be modeled using logistic regression

$$\begin{aligned} y_i &\sim \mathsf{Bernoulli}(\theta_i) \\ \theta_i &= \mathsf{logit}^{-1}(\eta_i) \\ \eta_i &= \beta_0 + \beta_{[\mathsf{trt[i],brand[i]],dose[i]]}} + \nu_{[\mathsf{site[i]]}} + \upsilon_{[\mathsf{siv[i]]}} + z_i^{\mathsf{T}} \alpha \end{aligned} \tag{1}$$

where

- $y_i$ is the binary outcome variable for participant $i$ having the value 1 when a mod/sev AE occurred and 0 otherwise
- $\theta_i$ is the probability of mod/sev AE in participant $i$
- $\eta_i$ is the log-odds of mod/sev AE in participant $i$
- $\beta_0$ is the mean log-odds of response in the population
- $\beta$ is a vector of parameters corresponding to variation from the mean log-odds of response in each brand, dose and treatment group
- $\nu$ is a vector of parameters corresponding to site effects
- $\upsilon$ is a vector of parameters corresponding to SIV type effects
- $\alpha$ is a vector of other covariates specified for model inclusions and $z_i^{\mathsf{T}}$ is a row from the design matrix for these covariates.

Each arm in the trial corresponds to a COVID-19 vaccination brand/dose pair coadministered with SIV or placebo. For convenience, we denote a treatment arm generically as $\mathsf{SIV}_j$ or $\mathsf{Placebo}_j$ where $\mathsf{SIV}$ and $\mathsf{Placebo}$ denote the intervention and control respectively and $j$ indicates a specific COVID-19 vaccination brand/dose pair. When the distinction between the SIV vs placebo treatment groups is unimportant (or we just want to refer to the COVID-19 vaccination brand/dose pair) then we will use $\mathsf{X}_j$.

Analysts have discretion to revise the parameterisation of the above model if needed. For example, in the initial configuration there is only a single site and therefore, there would be no need to include a site effect.

All variations and their rationale will be reported to the DSMC.

###### 3.5.1 Covariates

Age, sex, allergic history, use of prophylactic paracetamol and co-morbidities (specifically type-2 diabetes, hypertension and heart disease) and previous SARS-CoV-2 infection are believed to be associated with the occurrence of local and systematic reactions to COVID-19 vaccination (Ahamad, 2021; Folegatti, 2020; Li, 2021; Menni, 2021). Therefore, we envisage that some or all of these covariates will be included in the model, however, these covariates need to be finalised.

###### 3.5.2 Priors for primary model

Weakly informative priors will be used on all parameters. For the mean log-odds of response in the population $\beta_0$ we will use normal priors

$$\beta_0 \sim \mathsf{Normal}(0, 2.5^2) \tag{2}$$

For the effects characterising the variation from the mean log-odds of response in the population, sum-to-zero contraints will be imposed and $\beta$ will again use normal priors

$$\beta \sim \mathsf{Normal}(0, 2.5^2) \tag{3}$$

If additional sites start recruiting, we will model them as fixed effects for up to 3 sites and a random effect if more than 3 sites join. We will again use normal priors, specifying an exponential prior on the variance in the event that we implement as a random effect. All other terms will use normal priors

$$\beta \sim \mathsf{Normal}(0, 10^2) \tag{4}$$

Priors will be calibrated using the prior predictive distribution and can be modified subject to full disclosure and rationale.

#### 3.6 Secondary analyses

All bar two secondary outcomes are binary in their representation. For all secondary outcomes with a binary representation we will use logistic regression with linear predictor as specified in the Analysis of primary outcome section. The linear predictor will be extended as necessary to include, for example, terms to account for study visit.

##### 3.6.1 Ordinal outcomes

Outcomes `SS1` and `SS2` are ordinal with levels

- none
- mild
- moderate
- severe and
- life-threatening

These will be analysed using a Bayesian cumulative logistic model adjusted for relevant prognostic and stratifying factors. The model will assume proportional effects across the ordinal scale.

##### 3.6.2 Priors for secondary models

We will adopt priors equivalent to those specified for the primary analysis.

#### 3.7 Safety outcomes

Safety analyses beyond those implicit in the primary and secondary outcomes are not defined for FluVID.

#### 3.8 Subgroup analyses

Subgroup analyses will examine treatment effect heterogeneity by introducing interaction terms between prognostic covariates and the treatment term.

##### 3.9 Sensitivity analyses

Sensitivity will explore effects within contemporaneous cohorts of patients (i.e. by interim analysis). The effects of variation in prior specification will also be explored, for example fitting models via maximum likelihood.

##### 3.10 Missing data

While no imputation of the response will be incorporated into the analyses, we will examine worst and best-case scenarios. For example, as a worst case, assume all missing data on the interventional arms result in the occurrence of mod/sev AE and all missing data on the control arms do not.

##### 3.11 Model deviations

The analysis models will be assessed for adequacy and additional models (either simpler or more complex) may be investigated as part of checks of sensitivity, stability, and model fit. If any issues or concerns arise (for example, strong evidence of interactions across treatment domains), all changes or updates to the specified primary model will be documented and reported.

##### 3.12 Software

Data processing will be performed using R. Models will be fit in R using Stan via the `rstan` or `cmdstanr` packages.

### 4 Decision Procedures

Formally, the decision procedures fall under the remit of trial governance, but we include a high level summary here for convenience. For adaptations internal to the trial, predefined rules are evaluated based on statistical decision quantities derived from the primary model and pre-specified thresholds, see Trial Decision Criteria and Trial Adaptation. When a decision threshold is exceeded and criterion is met, the DSMC will consider the results and the recommended actions, which may or may not include public reporting. The DSMC may decide to present that recommendation to the Trial Steering Committee (TSC) who are then responsible for executing the recommendation.

The following simplified steps are typically involved in the process (this list is purely indicative and not intended to be a complete enumeration of the possibilities):

1. interim analysis results in a criterion being met, see Trial Decision Criteria and Trial Adaptation
2. results reported to DSMC for review, see Reporting
3. DSMC report that a criterion has been met and may pass on recommendations to the TSC
4. all participants enrolled up until the time when the action was enacted are followed up
5. a final analysis is run on the completed follow-up for those enrolled participants, see Final analyses
6. the final analysis is used as the basis for further reporting, see Reporting

### 5 *Trial Decision Criteria*

Trial decisions evaluated at the interim analyses will be informed based on the following definitions derived from the joint posterior distribution associated with the primary analysis. These definitions (and others) will also be used within the context of reporting both primary and secondary outcomes at the final analysis.

#### 5.1 Inferiority

Using the notation introduced under the Analysis of primary outcome section, the difference in the probability of a mod/sev AE when coadministered with a COVID-19 vaccination brand/dose pair and SIV relative to coadministration of the same COVID-19 vaccination brand/dose pair with placebo is

$$\delta_j = \theta_{[\mathsf{SIV},j]} - \theta_{[\mathsf{Placebo},j]} \tag{5}$$

and we define inferiority as the posterior probability that this difference exceeds zero

$$\phi_j = \mathsf{Pr}(\delta_j > 0) \tag{6}$$

where $\phi_j$ denotes the probability of inferiority and $j$ denotes a particular COVID-19 vaccination brand/dose pair.

At each analysis, the probability of inferiority will be compared to a threshold of 0.985. If this threshold is exceeded then a trial decision of inferiority will be made for the COVID-19 vaccination brand/dose pair under consideration.

Table 3: Intervention inferiority

| Decision | Comparison | Quantity | Threshold | Action |
|---|---|---|---|---|
| $\mathsf{SIV}_j$ is inferior | $\mathsf{SIV}_j$ vs $\mathsf{Placebo}_j$ | $\phi_j$ | $> 0.985$ | Stop enrollment into brand |

Note that under an inferiority signal, we recommend stopping all enrollment into both doses of the brand for which the decision was triggered.

#### 5.2 Non-inferiority

We define non-inferiority as the posterior probability that the difference characterised by $\delta_j$ is not greater than the non-inferiority margin of an absolute increase of $\Delta = 0.15$ on the probability scale.

$$\psi_j = \mathsf{Pr}(\delta_j < 0.15) \tag{7}$$

At each analysis, the probability of non-inferiority will be compared to a threshold of 0.985. If this threshold is exceeded then a trial decision of non-inferiority will be made for the COVID-19 vaccination brand/dose pair.

Table 4: Intervention non-inferiority

| Decision | Comparison | Quantity | Threshold | Action |
|---|---|---|---|---|
| $\mathsf{SIV}_j$ is non-inferior | $\mathsf{SIV}_j$ vs $\mathsf{Placebo}_j$ | $\psi_j$ | $> 0.985$ | Stop enrollment into brand/dose pair |

### 6 *Trial Adaptation*

As the trial proceeds, some aspects of the trial status may change, for example new sites may start recruiting or new COVID-19 vaccination brands may be introduced.

#### 6.1 Sequential Analyses

Interim analyses will be conducted frequently throughout the trial. The first analysis will be conducted after a minimum of 200 participants are enrolled to a single COVID-19 brand and have reached their 7-day follow up after visit 1. Invariably, there will be a delay between hitting an enrolment target and the data extract to allow for data cleaning and validation, during which time recruitment will continue.

Subsequent analyses will be scheduled at fixed intervals corresponding to every subsequent 100 enrolled participants reaching their 7-day follow up after visit 1 for as long as the trial is ongoing. If recruitment is slower or faster than expected, there may be either small or large changes in sample size from one analysis to the next, in which case the timing of analyses should be reviewed. The analyses will use all the data on participants who have reached the primary endpoint and have outcome data available to inform the current model. The results from these analyses will be used to direct progression of the trial.

#### 6.2 Early stopping

Recruitment into COVID-19 vaccination brand/dose pairs may be stopped early for sufficiently strong evidence of inferiority or non-inferiority, see Trial Decision Criteria.

- If a decision of intervention inferiority is made for a COVID-19 vaccination brand/dose pair then, following review, a trial conclusion is declared and the intervention recruitment will cease for that COVID-19 vaccination brand at both doses.
- If a decision of intervention non-inferiority is made for a COVID-19 vaccination brand/dose pair then, following review, a trial conclusion is declared and the intervention recruitment will cease for that COVID-19 vaccination brand/dose pair.

In some instances, the above actions may be delayed at the discretion of the DSMC.

#### 6.3 Adding COVID-19 vaccinations

It is highly likely that new brands of the COVID-19 vaccination will become available over time. Also, the current two-dose schedule may be revised to include additional doses. FluVID permits the introduction of new investigational arms, but requires a minimum enrollment of 100 participants into both the investigational and control arms for a COVID-19 vaccination brand/dose pair before trial decisions are permitted on that pair. Each new COVID-19 vaccination will require the preparation of a randomisation list, which will be implemented by a statistician blinded to the trial results, see Randomisation.

### 7 Reporting

Reporting covers considerations that relate to internal (e.g. reporting results of sequential analyses to DSMC) and external reporting (e.g. reporting for publication in the academic press). The analyses identified in this document will be included in future trial reports and manuscripts. Exploratory analyses not necessarily identified here may be performed to extend the planned analyses, but any unplanned analyses not specified here will be clearly identified in any statistical reports and manuscripts for publication.

#### 7.1 Reporting interim analyses

Interim analyses will be reported using a study-specific version of the Adaptive Health Intelligence DSMC reporting template (Totterdell and Jones, 2021). In brief, the report contains sections covering the following

- Executive summary
- Suggested communication to the study team investigators
- Enrolment status including rates of enrolment
- Protocol deviations
- Descriptive statistics for baseline demographic stratified by treatment arm
- CONSORT diagram of inclusions/exclusions
- Summary of data missingness
- Results from primary analysis
- Visualisation of results
- Results from safety analyses
- Line listing of adverse events

At a minimum, the results from the primary analysis will include:

- model parameter estimates summarised as means and quantiles (0.025, 0.5, 0.975)
- probabilities that each COVID-19 brand/dose pair is inferior
- probabilities that each COVID-19 brand/dose pair is non-inferior

#### 7.2 Reporting final analyses

There are currently no-specific considerations that relate to reporting final analyses beyond those raised in other sections.

#### 7.3 Reporting for publication

Public reporting of final analyses on brand/dose pairs may occur while the trial is ongoing.

### 8 *Appendices*

#### 8.1 CONSORT Template

Figure 1 provides a template to be used for reporting the progression of participants from enrollment through to analysis. The diagram will be prepared for the open and closed report at each interim and also for the final analysis.

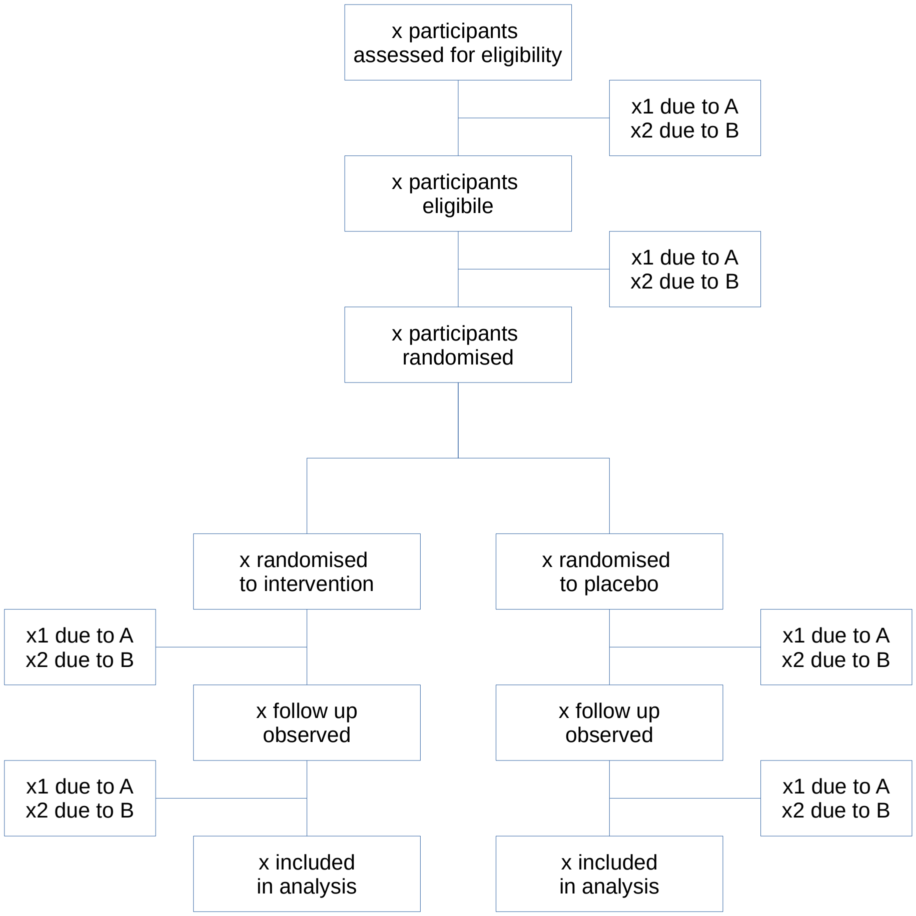

Figure 1: CONSORT template

##### 8.2 Data provision

Data cleaning including specification of any system of checks carried out by trial personnel to verify the integrity of the data are beyond the scope of this document.

The following provide examples of the main datasets required for the reporting to be completed. Data should be provided in third normalised form[1] (3NF). Standardised formats are to be used for fields. For example, dates are to be in `YYYY-mm-dd` format. Missingness is to be denoted by an empty field.

The following are purely indicative and a complete set of metadata for the required datasets is under development.

###### 8.2.1 Primary outcome data

Primary outcome data is required for all randomised participants regardless of whether the 7 day follow up has been reached. `ID` denotes the unique participant identifier. The second record is intentionally missing as part of the example.

Table 5: Primary outcome

| ID | FU | AE |
|---|---|---|
| M001 | 1 | 0 |
| M001 | 2 | |
| M002 | 1 | 1 |
| M002 | 2 | 1 |
| M003 | 1 | 1 |
| M003 | 2 | 1 |
| M004 | 1 | 0 |
| M004 | 2 | 1 |

Table 6: Allocation

| ID | Allocation |
|---|---|
| M001 | G0001 |
| M002 | G0002 |
| M003 | G0003 |
| M004 | G0004 |

Translation from allocation assignment identifier to treatment is held by the trial statistician.

###### 8.2.2 Participant data

The following participant data are required to report enrolment, progression through the trial and baseline characteristics. Missingness is intentional as part of example.

Table 7: Participant data

| ID | Enrolled | Randomised | Dose1 | Dose2 | Withdrawn | Completed | COVID | Dose | Age | Sex | Site |
|---|---|---|---|---|---|---|---|---|---|---|---|
| M001 | 2021-09-13 | 2021-09-13 | 2021-09-13 | 2021-09-21 | | 2021-10-20 | B1 | 1 | 29 | S1 | L1 |
| M002 | 2021-09-13 | 2021-09-13 | | 2021-09-21 | 2021-09-20 | 2021-10-20 | B1 | 2 | | S2 | L1 |

In line with 3NL, participant data with multiple entries should be reported as follows in which multiple comorbidities are reported per participant.

[1] Overview at https://www.guru99.com/database-normalization.html

Table 8: Participant comorbidity

| ID | Comorb |
|---|---|
| M001 | C2 |
| M001 | C1 |
| M001 | C4 |
| M002 | C1 |
| M002 | C2 |
| M003 | C4 |

Lookups are to be provided for all non-numeric fields that belong to a fixed set of options.

Table 9: Comorbidity lookup

| KeyComorb | Comorb |
|---|---|
| C1 | Cancer |
| C2 | Diabetes |
| C3 | CVD |
| C4 | other |

Table 10: Sex lookup

| KeySex | Sex |
|---|---|
| S1 | M |
| S2 | F |
| S3 | other |

Table 11: COVID-19 brand lookup

| KeyBrand | Brand |
|---|---|
| B1 | Pfizer |
| B2 | AZ |
